# Costs and health effects of CT perfusion-based selection for endovascular treatment of patients with a large vessel occlusion presenting within six hours after symptom onset A model-based health economic evaluation

**DOI:** 10.1101/2023.03.16.23287320

**Authors:** Henk van Voorst, Jan W. Hoving, Miou S. Koopman, Jasper D. Daems, Daan Peerlings, Erik Buskens, Hester F. Lingsma, Henk A. Marquering, Hugo W.A.M. de Jong, Olvert A. Berkhemer, Wim H. van Zwam, Marianne A.A. van Walderveen, Ido van den Wijngaard, Diederik W.J. Dippel, Albert J. Yoo, Bruce C.V. Campbell, Wolfgang G. Kunz, Charles B.L.M. Majoie, Bart J. Emmer, the CLEOPATRA, MR CLEAN Registry, CONTRAST consortium Investigators.

## Abstract

**Introduction:** Current stroke guidelines do not give uniform recommendations regarding the use of CT perfusion (CTP) for the selection of patients presenting within six hours after symptom onset for endovascular treatment (EVT). Model-based analyses can be used to estimate the potential long-term costs and health effects of CTP for patient selection.

**Methods:** In this nationwide retrospective cohort study with model-based health economic evaluation, 703 large vessel occlusion acute ischemic stroke patients with CTP imaging and EVT within six hours after symptom were included (Inclusion: January 2018–March 2022; trialsearch.who.int:NL7974). CTP-based EVT patient selection using varying ischemic core volumes (ICV) and core-penumbra mismatch ratios (MMR) was compared with providing EVT to all patients. Net monetary benefit (NMB) at a willingness to pay of €80,000 per quality-adjusted life year, the incremental cost-effectiveness ratio (ICER), the difference in costs (ΔCosts), and quality-adjusted life years (ΔQALY) per 1000 patients were the outcome measures.

**Results:** The cohort of patients with CTP and EVT used for simulations consisted of 391/703 males with a median age of 72 (IQR:62;81). Considering the most optimal ICV (≥110mL) and MMR (≤1.4) thresholds, CTP-based selection for EVT resulted in a loss of health (ΔQALYs: ICV-median:-3.3[IQR:-5.9;-1.1], MMR median:0.0 [IQR:-1.3;0.0]), limited additional costs or cost savings (ΔCosts: ICV-median:-€348,966[IQR:-€712,406;-€51,158], MMR-median:€266,336[IQR:€229,403;€380,095]), and an ICER and NMB with a wide IQR (ICER ICV-median:71,346[IQR:-16,517;181,241], MMR-median:312,955[IQR:-141,379;infinite]) (NMB ICV-median:€102,227[IQR:-€282,942;€431,923], MMR-median:-€278,850[IQR:-€457,097:-€229,403]).

**Conclusion:** In EVT-eligible patients presenting within six hours after symptom onset, excluding patients based on CTP parameters was not cost-effective and could potentially harm patients.

**Key points:** *What is already known on this topic:* Recent randomized clinical trials in patients with a large vessel occlusion and a large infarct region concluded that endovascular treatment (EVT) resulted in more favorable patient outcomes compared to best medical management. However, it remains largely unclear what the associated costs and health implications are in the long run of CT perfusion (CTP) based patient selection for EVT in patients presenting within six hours after symptom onset.

*What this study adds:* At optimized ischemic core volume (ICV) and core-penumbra mismatch ratio (MMR) thresholds, CTP-based selection for EVT resulted in a loss of health (ΔQALYs: ICV≥110mL median:-3.3[IQR:-5.9;-1.1], MMR≤1.4 median:0.0 [IQR:-1.3;0.0]) for similar costs (ΔCosts: ICV≥110mL median:-€348,966[IQR:-€712,406;-€51,158], MMR≤1.4 median:€266,336[IQR:€229,403;€380,095]) per 1,000 patients.

*How this study might affect research, practice or policy:* Selecting patients using CTP will likely result in a loss of health and at best a minor cost saving. Even in scenario’s considering unfeasibly low EVT benefit and in patients aged≥80 years CTP based patient selection for EVT was not cost-effective.

## Introduction

Acute ischemic stroke (AIS) patients with a large infarct region due to a large vessel occlusion (LVO) had a better outcome after endovascular treatment (EVT) compared to best medical management in three recent randomized clinical trials (RCTs).^1–3^ This is in contrast to earlier studies where no statistically significant benefit of EVT was found in patients with a large infarct who present within six hours after symptom onset.^4^ Furthermore, the benefit of EVT was higher in RCTs that used CTP measures such as ischemic core volume (ICV) and core-penumbra mismatch ratio (MMR) as an inclusion criteria.^4^ Additionally, the benefit of EVT declines depending on ICV in patients aged ≥75 years.^5^ Since recent large stroke trials had restrictive inclusion criteria, varying imaging protocols, and only evaluated patient outcome at 90 days follow-up,^1–3^ it remains partially unclear what the long-term costs and health effects are of selecting patients for EVT based on CTP measures. Health economic model-based analyses could be used to estimate long-term costs and health effects for various scenarios and uses of CTP for EVT patient selection.

Three previous model-based studies had conflicting results regarding the cost-effectiveness of CTP for patients presenting within six hours.^6–8^ These three studies were based on fixed assumptions for the EVT effect, EVT effect modification due to CTP measures, and fixed decision threshold based on CTP measures. As a result, these studies do not consider population variations and uncertainty in current evidence.^9,10^ Namely, the EVT effect could be lower in patients with a large infarct or in elderly subgroups.^5^ In addition, the optimal ICV or MMR thresholds for EVT selection remain a matter of debate. Furthermore, due to improvements in CTP acquisition and analytical software, previous evidence for ICV and MMR-based EVT effect modification and associations with functional outcome might be inaccurate.^11^ Finally, costs and health effect estimates from previous studies were adapted from a US perspective which might not apply to European and other healthcare systems.^6–8^

We aimed to quantify the long-term costs and health effects of CTP-estimated ICV and MMR to select patients within six hours after stroke onset for EVT in the Netherlands compared to providing EVT to all patients. Furthermore, we aimed to identify scenarios where CTP-based patient selection for EVT might be cost-effective.

## Methods

### Study design

The Cost-effectiveness of CT perfusion for patients with acute ischemic stroke (CLEOPATRA) is a Dutch nationwide, retrospective, multicenter health economic modeling study using patient-level data and literature parameters.^12^ A cohort of patients with an LVO that received EVT after CTP imaging with 90-day functional outcome according to the modified Rankin Scale (mRS) was considered. During inclusion, the standard of care was to provide EVT to all patients regardless of the CTP measures.^13^ We compared CTP-based patient selection for EVT (“CTP select EVT” arm) with providing EVT to all patients (“all EVT” arm). The decision to offer or withhold EVT in the CTP arm was dependent on an ICV or MMR threshold per patient. Since we used observational data of patients who underwent EVT, available ORs for treatment effect, ^9,14^ and treatment effect modification due to CTP measures^10^ were used to generate the 90-day mRS as if those patients did not undergo EVT (referred to as the ‘no EVT group’). We assumed that EVT could directly follow after imaging and that CTP would not delay time to EVT. The methodology for the no EVT group generation is described in **Online Supplement A**.

### Data collection

Patients who had CTP before EVT in an EVT-capable stroke center for an anterior circulation LVO between January 2018 and March 2022 were included from the following data sources: the Multicenter Randomized Clinical Trial of Endovascular Treatment of Acute Ischemic Stroke in The Netherlands (MR CLEAN)-NO IV (ISRCTN80619088),^15^ MR CLEAN-MED (ISRCTN76741621),^16^ the MR CLEAN Registry,^17^ and a local cohort of patients from our comprehensive stroke center (Amsterdam UMC location University of Amsterdam, Amsterdam, The Netherlands). A detailed overview of the study-specific inclusion and exclusion criteria were previously published together with predefined modeling procedures and study endpoints.^12^ **Online Supplement B** describes minor changes to the protocol.^12^ CTP acquisition, post-processing, and quality assessment are presented in **Online Supplement C**. ^12^ For patients included in the MR CLEAN-MED and MR CLEAN-NO IV trials, deferred consent was received, for patients included in the MR CLEAN Registry and the retrospective local cohort a waiver for informed consent was provided by the institutional review board. Data is available upon reasonable request and following local privacy regulations.

### Modeling approach

A Markov model with patient-level micro-simulations was used to simulate 5- and 10-year follow-up. The model consisted of a short-term 90-day post-AIS model to simulate EVT patient selection strategies followed by a long-term yearly model (**Figure 1**) and was used to simulate functional outcome measured with the mRS. In the long-term model, deterioration of mRS was simulated based on the probability of stroke recurrence^18^ and death^19^ inflated with patient-specific Hazard Ratios (HR).^20^ Simulations and analyses were performed with publicly available custom-developed Python software (github.com/anonymous/CLEOPATRA). All model input parameters are described in **Online Supplement D**.

**Figure 1:**
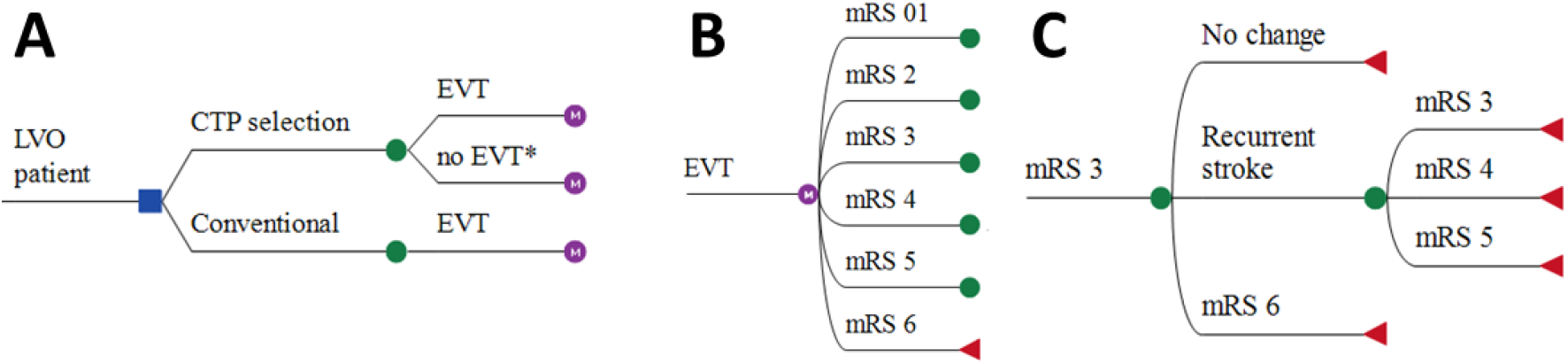
Markov model structure. A) Patients with a confirmed large vessel occlusion (LVO) who were eligible for endovascular treatment (EVT) were simulated. Each patient was simulated as if NCCT and CTA imaging was used with or without additional CT perfusion (CTP) for selection of patients for EVT. In the CTP arm, the decision for EVT was based on varying thresholds for ischemic core volume and core-penumbra mismatch ratio. In the conventional arm, all patients underwent EVT. B) after EVT or no EVT the 90-day modified Rankin Scale (mRS) was modelled based on observed mRS (EVT group) or adjusted mRS with ORs (no EVT group). C) After the 90-day outcome, yearly mRS transitions were modelled based on death and recurrent stroke rates. *: The proportion of patients that do not receive EVT differs based on the set ischemic core volume or core-penumbra mismatch ratio thresholds.

### Costs and QALYs

Costs in euros (€) from a healthcare payer perspective and QALYs over the simulated period were derived per mRS sub-score in a previous study^21^ and predefined in our protocol.^12^ **Online supplement D** describes the acute care and long-term follow-up costs. Acute care costs included personnel cost, radiological imaging, EVT procedure, and thrombolysis costs. Long-term follow-up costs considered In-hospital care use, outpatient clinic visits, rehabilitation, formal homecare, and long-term institutionalized care. Present values of simulated costs and QALYs were calculated based on a discounting rate of 4% for QALYs and 1.5 % for costs yearly.^22^ Historical and forecasted inflation rates for healthcare costs were used to adjust the costs over time.^23,24^

### Outcome measures

The primary outcome was the net monetary benefit (NMB: Formula 1) at a willingness to pay (WTP) of €80,000 per QALY of CTP-based EVT patient selection (CTP) compared with the conventional imaging arm (no CTP). Secondary outcome measures were the incremental cost-effectiveness ratio (ICER: Formula 2), and differences in cost (ΔCosts) and quality-adjusted life-years (ΔQALYs) between the simulated CTP and no CTP arms.

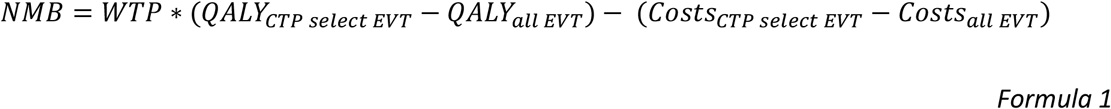

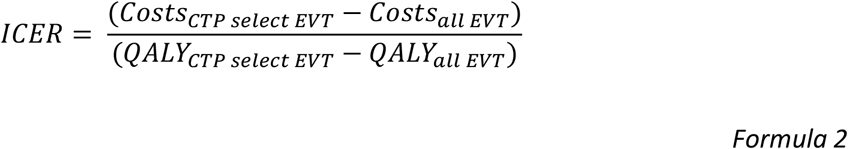

### Baseline and sensitivity analyses

Mean values of the model inputs were used for the baseline simulation including all patients once. Baseline simulation EVT patient exclusion thresholds were ≥70mL for ICV and ≤1.8 for MMR. In the one-way sensitivity analyses, the effect on outcomes of a 10% increase or decrease of input parameters relative to the baseline simulation was simulated. In the probabilistic sensitivity analyses (PSAs), 1,000 cohorts of 703 patients were sampled with replacement from the original data. Each cohort contains a proportion of patients that did not receive EVT in the CTP arm since they had an ICV above or MMR below the decision threshold. ICV and MMR values were varied with increments of 10 between 0 and 150 mL and increments of 0.2 between 1.4 and 2.4, The decision threshold with the highest median NMB across all cohorts was chosen as the optimal setting. Per simulated cohort, the ΔCosts and ΔQALYs between the CTP and no CTP arms were computed. All PSA results were reported as the median with interquartile range (IQR) over the 1,000 simulated cohort per 1,000 patients.

A baseline PSA was conducted using an OR for the EVT effect from the MR CLEAN trial (OR:1.67[95%CI:1.21-2.30]).^14^ We performed dedicated PSAs altering the OR and 95%CI for EVT effect with -0.5, -0.3, +0.3, and +0.82. The lower bound (−0.5) corresponds to the EVT effect in patients with a large infarct in NCCT (Alberta Stroke Program Early CT [ASPECTS] score≤5) and the upper bound (+0.82) refers to the pooled EVT effect in the HERMES meta-analysis.^9^ The regression analyses for EVT effect modification due to CTP estimated ICV and MMR by Campbell et al. were repeated as the initial work only reported p-values (full analyses in **Online supplement D**).^10^ An OR for EVT effect modification due to ICV (OR:0.98 [95%CI:0.881-1.091] per 10 mL) and MMR (OR:1.010[95%CI:0.994;1.026]) per one point change) were used for baseline simulations. Since these ORs were based on CTPs from trials published between 2015 and 2017 with limited brain coverage,^10,25^ the ORs and 95%CIs for effect modification were altered by -0.1 and -0.05 for ICV, and +0.1 and +0.05 for MMR to consider technological improvements in CTP software. Finally, we simulated the most favorable CTP-based EVT patient selection PSA scenario considering patients aged ≥80 years, a 0.3 decrease of the EVT effect OR (OR:1.37[95%CI:0.91-2.0]), and a 0.1 decrease of the ICV or MMR based EVT effect modification (OR:0.88[95%CI:0.781-0.991]).

## Results

### Descriptive statistics

A total of 703/1,122 patients from the CLEOPATRA database were considered in this study. **Online Supplement E** contains the exclusion reasons (**Figure E1**) and tables with baseline characteristics (**Tables E1-3**). The main exclusion reasons were: presentation beyond six hours after symptom onset and CTP source data that could not be processed due to anonymization at local sites or discrepancies between local hardware and our CTP software. There were 70/703(10.0%) patients with an ICV≥70mL, 23/703(3.3%) patients with an ICV≥110mL, 19/703(2.7%) patients with an MMR≤1.8, and 5/703(0.7%) patients with an MMR≤1.4.

### Baseline model and one-way sensitivity

In the baseline simulations, excluding patients with an ICV≥70mL for EVT resulted in a loss of health (ΔQALY:-33.8), higher costs (Δcosts:€385,866), an ICER of -11,432, and an NMB of -€3,086,208 per 1,000 patients. Excluding patients with an MMR≤1.8 for EVT resulted in a loss of health (ΔQALY:-5.8), higher costs (Δcosts:€54,836), an ICER of -9,834, and an NMB of -€500,913 per 1,000 patients. **Figure F1** (**Online Supplement F**) describes the ten model parameters that affected the NMB the most in strategies using ICV or MMR for EVT patient selection. For the ICV simulations, the QALYs for mRS 0-2, long-term costs in the third year and beyond for mRS 5, and costs of EVT affected NMB the most. For the MMR simulations, the QALYs in mRS 3-4, costs of EVT, and the costs related to mRS 4 in the first year after AIS affected NMB the most.

### Probabilistic sensitivity analysis

**Figure 2** depicts the PSA results in incremental cost-effectiveness ratio plots (**Figures 2A-B**), and the NMB per decision threshold for ICV and MMR-based patient selection (**Figures 2C-D**). A more detailed overview of the results is provided in Online Supplement F. For all PSAs the optimal thresholds for EVT patient exclusion using were ICV≥110 mL and MMR≤1.4. Using ICV to exclude patients for EVT resulted in a loss of health (ΔQALYs 5-year median:-3.3[IQR:-5.9;-1.1], 10-year median:-4.3[IQR:-9.1;-0.7]), a limited cost saving (ΔCosts 5-year median: -€348,966[IQR:-€712,406;-€51,158], 10-year median:-€246,679[IQR:-€733,714;€191,840]), a wide IQR for the ICER (ICER 5-year median:71,346[IQR:-16,517;181,241], 10-year median:31,244[IQR:-48,780;141,749]), and an NMB close to zero (NMB 5-year median:€102,227[IQR:-€282,942;€431,923], 10-year median: -€97,781 [IQR:-€644,889;€420,136] per 1,000 patients. Using MMR to exclude patients for EVT resulted in a limited to no loss of health (ΔQALYs 5-year follow-up median:0.0[IQR:-1.3;-0.0], 10-year follow-up median:0.0[IQR:-1.6;-0.0]), higher costs (ΔCosts 5-year follow-up median:€266,513[IQR:€229,403;€380,110], 10-year follow-up median:€279,661[IQR:€229,403;€464,756]), a wide IQR for the ICER (ICER 5-year follow-up median: 312,955[IQR:-141,379;infinite], 10-year follow-up median:249,523[IQR:-118325;infinite]), and a negative NMB (NMB 5-year follow-up median:-€278,626[IQR:-€456,559;-€229,403], 10-year follow-up median:-€279,614 [IQR:-€555,168;-€228,836]) per 1,000 patients. Due to the limited number of patients with an ICV above (≥110mL) and MMR below (≤1.4) the optimal decision threshold for EVT patient exclusion, the ΔQALY results were low. This effect was more profound for MMR compared to ICV-based patient selection; low MMR values were less common than high ICV values. The effect on costs in this small group of patients excluded for EVT was due to a decrease in long-term care costs, related to earlier death, and due to the absence of EVT costs. Subsequently, low ΔQALY values resulted in an ICER with a wide IQR.

### Changes in EVT effect and ICV or MMR-based effect modification

Table 1 describes the ICERs for different shifted values for the EVT effect and EVT effect modification due to ICV or MMR from the baseline levels. Online Supplement F contains more scenarios and reports the ICER, ΔCosts, and ΔQALYs. For the simulations using ICV, a higher EVT effect resulted in a higher cost saving, a higher QALY loss, and a higher ICER and NMB. A decrease in the ICV-based EVT effect modification resulted in a higher cost saving, a higher QALY loss, with a higher ICER and NMB. For the simulations using MMR, changes in EVT effect or the MMR-based EVT effect modification had a negligible effect on the outcome measures. This was mainly due to the small proportion of patients that had an MMR≤1.4. Even at the lowest EVT effect and ICV or MMR-based EVT effect modification (lower left corner in the table), the IQR of ICER and NMB included zero indicating no benefit of the use of CTP-based patient selection.

**Table 1.** NMB for alternative scenarios of EVT effect and effect modification due to ICV and MMR per 1000 patients. Results are presented as median with IQR, the baseline follow-up horizon of 5-year was used. The optimal ICV threshold for patient exclusion for EVT was in all scenarios ≥110mL, the optimal MMR threshold for patient exclusion for EVT was ≤1.4 in all scenarios. Optimal thresholds were based on the maximum net monetary benefit at a willingness to pay of €80,000. *: The baseline ORs from the MR CLEAN trial for EVT effect (9) and the study by Campbell et al. for ICV based effect modification (4). **: Just below the mean EVT effect of patients with a large infarct on NCCT (ASPECTS<6) in the HERMES pooling. ***: EVT effect found in the HERMES pooling (1). A lower EVT effect and a lower ICV or MMR based EVT effect modification are more beneficial scenarios for using CTP for EVT patient selection. ICER: incremental cost effectiveness ratio. ICV: ischaemic core volume. MMR: core-penumbra mismatch ratio. EVT: endovascular treatment. OR: odds ratio. CTP: CT perfusion.

| CTP measure and optimized threshold | Follow-up horizon (years) | Effect modification due to ICV or MMR (OR) | Net monetary benefit (NMB) in € |  |  |  |  |
| --- | --- | --- | --- | --- | --- | --- | --- |
|  |  |  | EVT effect OR |  |  |  |  |
|  |  |  | 1.17<br>(95%CI:0.71;1.8)** | 1.37<br>(95%CI:0.91;2.0)<br>) | 1.67<br>(95%CI:1.21;2.3)<br>* | 1.97<br>(95%CI:1.51;2.6) | 2.49<br>(95%CI:2.03;3.12)*<br>** |
| ICV≥110mL | 5 | 0.98 (95%CI: 0.881;1.091)* | 75,260<br>(-256,606;356,220) | 92,540<br>(-274,492;387,168) | 102,227<br>(-282,942;431,923) | 115,014<br>(-282,947;472,304) | 126,436<br>(-294,179;516,591) |
|  | 5 | 0.93 (95%CI: 0.831;1.041) | 52,532<br>(-158,294;247,178) | 58,740<br>(-171,751;270,871) | 71,360<br>(-189,586;298,635) | 81,540<br>(-202,099;334,318) | 95,139<br>(-235,410;378,627) |
|  | 5 | 0.88 (95%CI: 0.781;0.991) | 44,361<br>(-84,207;180,651) | 45,137<br>(-97,373;189,920) | 52,380<br>(-117,014;211,993) | 57,805<br>(-131,228;233,197) | 69,596<br>(-146,628;265,746) |
|  | 10 | 0.98 (95%CI: 0.881;1.091)* | -119,896<br>(-587,168;333,612) | -112,504<br>(-620,679;366,957) | -97,781<br>(-644,889;420,136) | -103,484<br>(-659,676;460,891) | -74,712<br>(-685,404;519,196) |
|  | 10 | 0.93 (95%CI: 0.831;1.041) | -82,733<br>(-430,820;215,513) | -89,848<br>(-460,125;247,493) | -95,305<br>(-508,231;268,169) | -94,125<br>(-521,341;297,295) | -96,801<br>(-565,620;347,538) |
|  | 10 | 0.88 (95%CI: 0.781;0.991) | -24,078<br>(-262,408;153,870) | -38,411<br>(-295,091;163,770) | -47,193<br>(-327,253;184,168) | -56,497<br>(-369,169;205,562) | -69,079<br>(-415,466;234,382) |
| MMR≤1.4 | 5 | 1.010 (95%CI: 0.994;1.560) | -263,889 (-433,414;-222,141) | -267,393 (-443,666;-225,803) | -278,626 (-456,559;-229,403) | -279,614 (-465,361;-229,379) | -279,614 (-477,472;-229,403) |
|  | 5 | 1.060 (95%CI: 1.044;1.610) | -264,070 (-433,655;-222,185) | -267,548 (-444,090;-225,943) | -278,740 (-456,835;-229,403) | -279,614 (-465,472;-229,360) | -279,614 (-477,681;-229,403) |
|  | 5 | 1.110 (95%CI: 1.094;1.660) | -264,073 (-433,887;-222,227) | -267,696 (-444,428;-226,076) | -278,850 (-457,097;-229,403) | -279,614 (-465,577;-229,342) | -279,614 (-477,880;-229,403) |
|  | 10 | 1.010 (95%CI: 0.994;1.560) | -271,910 (-520,757;-219,184) | -279,614 (-534,853;-222,380) | -279,614 (-555,168;-228,836) | -279,614 (-570,464;-229,403) | -279,614 (-589,096;-229,403) |
|  | 10 | 1.060 (95%CI: 1.044;1.610) | -272,133 (-521,893;-219,316) | -279,614 (-535,771;-222,362) | -279,614 (-555,982;-228,996) | -279,614 (-571,036;-229,403) | -279,614 (-589,859;-229,403) |
|  | 10 | 1.110 (95%CI: 1.094;1.660) | -272,027 (-521,338;-219,251) | -279,614 (-535,334;-222,371) | -279,614 (-555,585;-228,917) | -279,614 (-570,770;-229,403) | -279,614 (-589,487;-229,403) |

**Figure 2.**
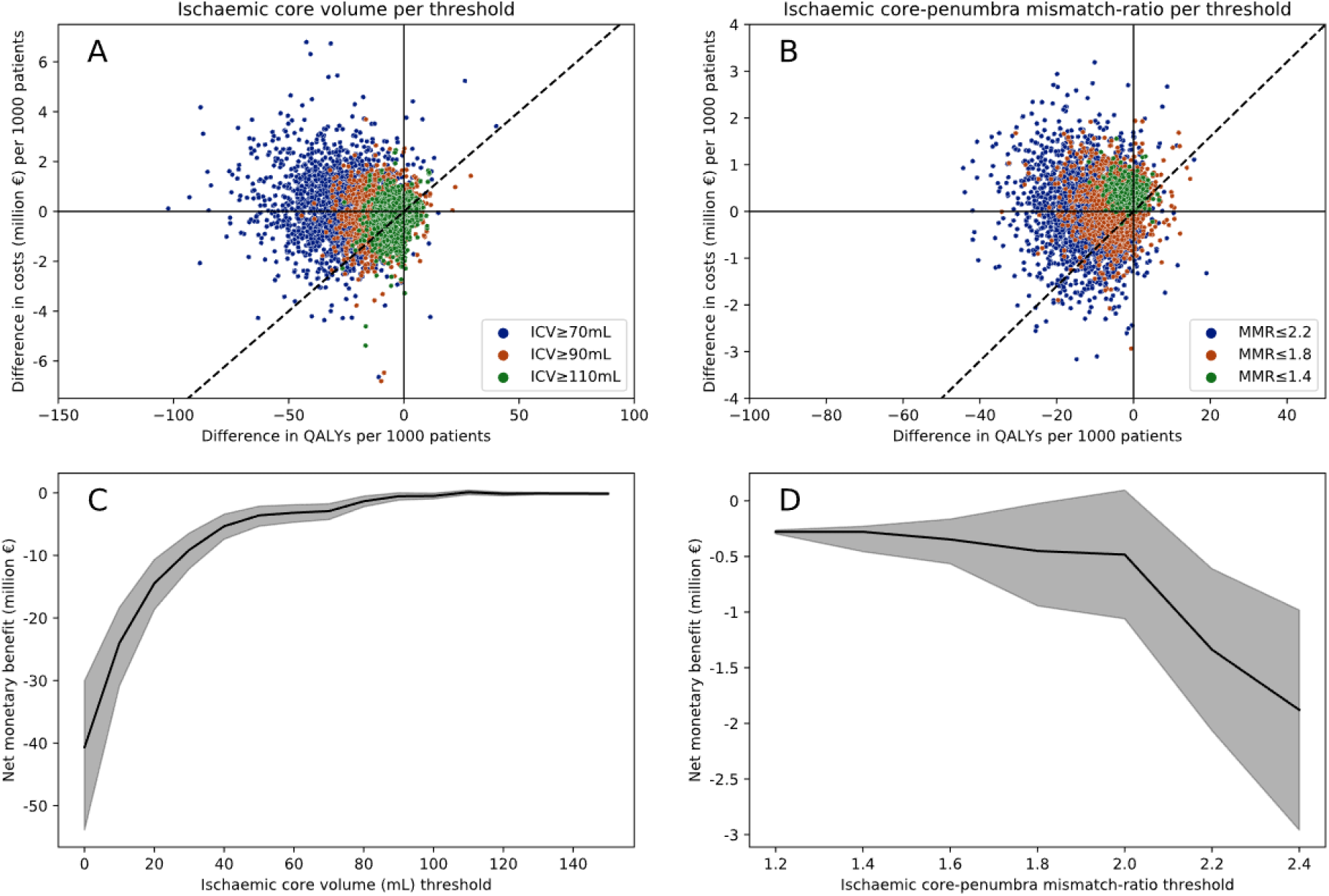
Probabilistic sensitivity analyses results. 4A-B: Each point in the scatterplot is a cohort of patients that was simulated twice; one time as if the optimal CTP measure (CTP arm) was used to exclude patients for EVT and one time as if all patients received EVT (no CTP arm). The x and y coordinates of this cohort are based on the differences in QALYs and costs between these two simulated arms and thus represent the benefit or harm due to CTP-based patient selection. The dashed diagonal line represents the willingness to pay of €80,000 per QALY; below the line is cost-effective. The colors were used to show the effect of different ICV and MMR decision thresholds for patient exclusion for EVT. 4C-D: The trend observed in 4A-B for different decision thresholds is presented per 10mL ICV and 0.2 MMR decision threshold increments on the x-axis with a net monetary benefit on the y-axis. The black line represents the median, the grey area the inter-quartile range. The net monetary benefit aggregates the QALY and cost differences between the CTP and no CTP arms to a single value by multiplying the QALY difference with €80,000 and subtracting the cost difference. 4A: Incremental cost-effectiveness ratio plot for varying ICV thresholds for EVT patient selection. 4B: Incremental cost-effectiveness ratio plot for varying MMR thresholds for EVT patient selection. 4C: The net monetary benefit for each 10 mL increment of the ICV threshold, the optimal threshold was ≥110mL. 4D: Similar to 4C but considers the MMR, the optimal threshold was ≤1.4. QALYs: quality-adjusted life-years. ICV: ischemic core volume. MMR: core-penumbra mismatch ratio. CTP: CT perfusion.

### Elderly subgroup: most favorable CTP for EVT selection scenario

The reduced life expectancy in the subgroup of patients ≥80 years resulted in a less profound loss of QALYs (ΔQALYs ICV median:0.0[IQR:-0.8;0.5], MMR median:-1.2[IQR:-8.8;0.0]) and additional costs (ΔCosts ICV median:€34920[IQR:-€293,371;€524,269], MMR median:-€229,448[IQR:-€1,178,568;€461,962]). However, IQRs of the ICER (ICV median: 120,757[IQR:-343,877;932,702], MMR median:88,483[IQR:-61,501;65,3609]) and NMB (ICV median:€23,425[IQR:-€461,957;€305,633], MMR median:€108,270[IQR:-€775,446;€1,004,928]) per 1,000 patients still included zero. **Figure 3** contains the ICER plot for the elderly subgroup PSA with the most favorable scenario for ICV and MMR based patient selection.

**Figure 3.**
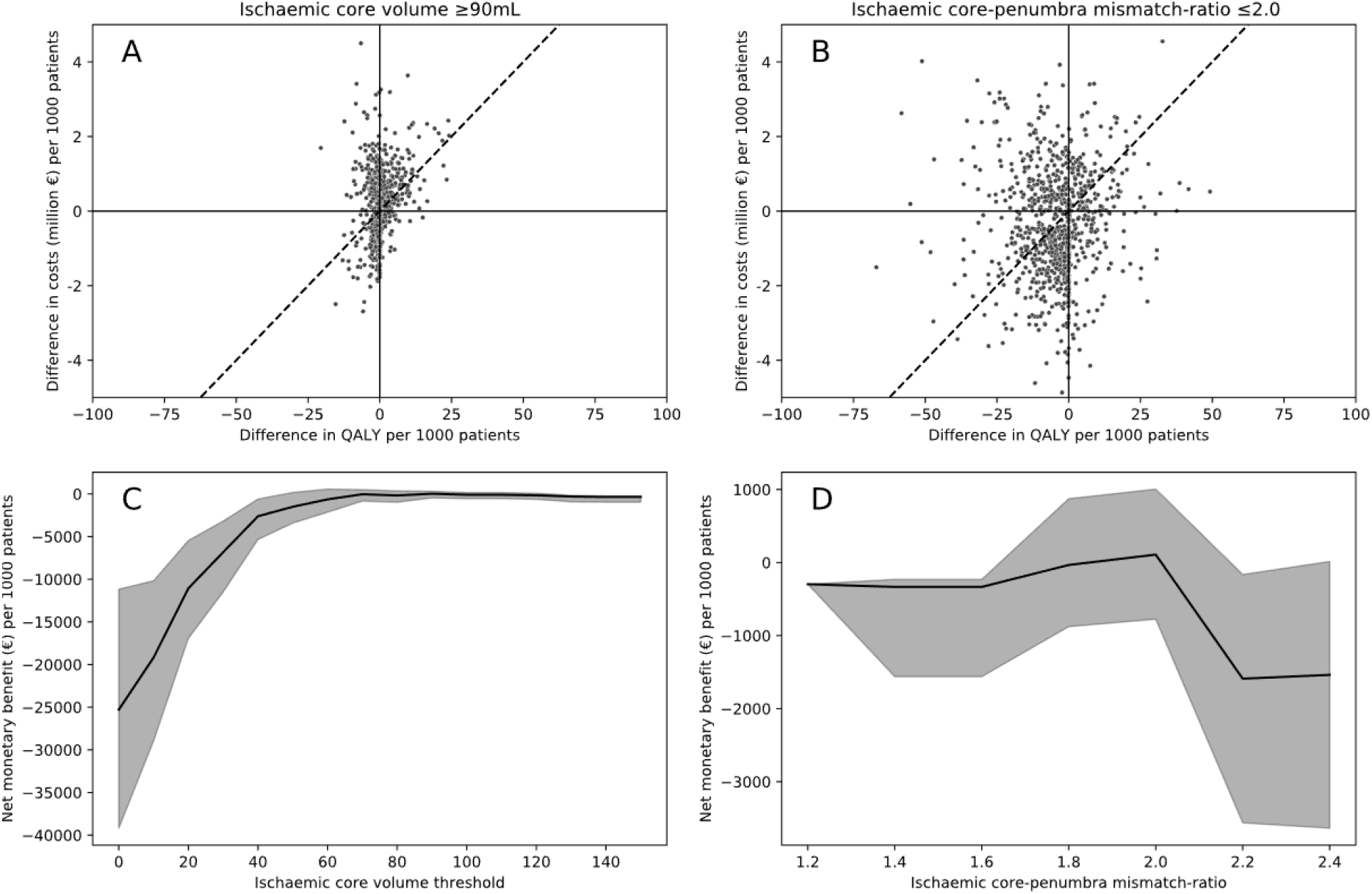
ICER plot for CTP based EVT selection in an elderly population. For this simulation a follow-up horizon of 10 years, 0.3 decrease of the EVT effect (OR:1.37[95%CI:0.91-2.0]), and a 0.1 decrease of the ICV or MMR based EVT effect modification (OR:0.88[95%CI:0.781-0.991]) were used to simulate cohorts with patients aged ≥80 years. This is a favorable but unlikely scenario for CTP-based selection for EVT. 5A: PSA performed for ICV with an optimal threshold of ≥90mL for excluding patients for EVT. 5B: PSA performed for MMR with an optimal threshold of ≤2.0 for excluding patients for EVT. 5C-D: per ischemic core volume and mismatch-ratio threshold the NMB for the most favorable scenario for CTP-based EVT patient selection. EVT: endovascular treatment. ICV: ischemic core volume. MMR: core-penumbra mismatch ratio. PSA: probabilistic sensitivity analysis.

## Discussion

In the baseline scenario, using the baseline CTP parameter estimates ICV or MMR for EVT patient selection within six hours after symptom onset resulted in no additional costs or cost savings, a loss of health, an NMB close to zero, and an ICER with a wide IQR compared to providing EVT to all patients. We observed a reduction in the loss of health and an increase in costs if the EVT benefit would be lower and when the benefit of EVT diminishes further for patients with a larger ICV and a lower MMR. For a low EVT effect and high reduction of EVT effect modification due to ICV and MMR, the simulation resulted – at best – in a negligible loss of health for no additional costs and thus an NMB close to zero. Even under unlikely favorable conditions, in patients aged ≥80 years, assuming a low EVT benefit, and a high decline of EVT effect due to ICV or MMR, CTP-based EVT patient selection was not cost-effective.

To interpret the results of this study, the presented scenarios should be compared with the best available evidence. The EVT effect and EVT effect modification due to ICV and MMR were more conservative compared to the three recent EVT trials.^1–3^ Therefore, the results from this study at baseline simulation values might present a scenario less favorable for outcomes after EVT and thus more favorable for CTP-based patient selection. In light of recent and ongoing AIS workflow improvements, it seems likely that the benefit of EVT will only increase in the coming years.^26^ Since we did not observe a benefit of CTP-based patient selection in more favorable scenarios, technological improvements of CTP acquisition and software analyses might be futile to further improve CTP-based EVT patient selection.^10^ Finally, in the most favorable CTP-based EVT patient selection scenario, we considered a lower EVT effect for patients aged ≥80 years. However, current evidence suggests a higher EVT effect for patients aged ≥85 years while we used a reduced EVT effect.^27^ Other research suggests that patients aged ≥75 years with ICV ≥50 or ≥85 mL might have a lower EVT effect.^5^

Our study has limitations. First, we did not use prospectively collected data regarding treatment effect and CTP-based effect modification, long-term mRS follow-up, and cost data. Due to the retrospective nature of this study, we could have missed patients that would not benefit from EVT such as patients with a worse pre-morbid functional status. Second, we used cost data from a healthcare payer perspective, neglecting potential effects due to indirect costs related to for example labor productivity in young AIS patients. Third, CTP might be used to improve occlusion detection and IVT administration.^9,28^ Thus, the actual added value of CTP in the diagnostic workup of patients with AIS might be higher due to the beneficial effect on occlusion detection. Fourth, we only included CTP results from one single vendor with fixed settings. This reduces part of the variation in measurements compared to real-world measurements which likely vary between centers due to the variation of analytical CTP software packages.^29,30^ As a result – even though no benefit of CTP for patient selection for EVT was found – findings from this study may be too optimistic, making it very unlikely that using CTP for the selection for EVT within 6 hours after symptom onset is currently – or, with improvements, will soon become – cost-effective. Further improvements in the CTP acquisition and analysis could also alter the association between the CTP measures and outcome, without EVT effect modification, and thus affect the cost-effectiveness. This effect is not covered in this study.

Future work should investigate the cost-effectiveness of CTP-based patient selection in sub-populations with a high risk of poor functional outcome. For example, patients presenting after six hours from stroke onset, elderly patients, patients with pre-morbid disability, or complex co-morbidities as these patients might have a reduced benefit from EVT.^31^ However, as represented by the baseline characteristics in this study, this combination of highly unfavorable baseline characteristics is rare in our AIS population.^5^ Furthermore, long-term follow-up studies and cost-effectiveness analyses of the large infarct RCTs will provide a higher level of evidence.^1–3^

## Conclusion

In this Dutch, nationwide cohort study with a model-based health economic evaluation, the use of CTP-estimated ischemic core volume or core-penumbra mismatch ratio to exclude patients presenting within six hours after symptom onset from endovascular treatment resulted in a loss of health and no cost savings. Additional simulations revealed that under unlikely favorable conditions, these findings did not change. The use of CTP-based parameters to exclude patients from EVT is unlikely to be cost-effective and may potentially harm EVT-eligible patients who present within six hours after onset.

## Data Availability

Data sharing statement
The complete de-identified patient datasets from the MR CLEAN-NO IV, and MR CLEAN-MED trials will be available from 18 months after publication until 15 years from publication. Data can be obtained from https://www.contrast-consortium.nl/data-request-form/. The data will be made available to researchers who are CONTRAST consortium members or collaborators, and whose proposed use of the data has been approved by the CONTRAST data access and writing committee. The data will be made available for specified purposes, as defined in the sub study proposal and approved by the CONTRAST data access and writing committee. The data will be made available after approval of the proposal by the CONTRAST data access and writing committee. To ensure publication transparency and quality, researchers should adhere to the CONTRAST publication policy, accessible on https://www.contrast-consortium.nl/publication- policy-contrast/. For the patients included in the MR CLEAN Registry and the local cohort, individual patient data cannot be made available under Dutch law since we did not obtain patient approval for sharing individual patient data. All syntax files and output of statistical analyses are available on reasonable request to the corresponding author.

https://www.contrast-consortium.nl/

https://www.mrclean-trial.org/

## Abbreviations

QALY: Quality-adjusted life year
NMB: Net monetary benefit
ICER: Incremental cost-effectiveness ratio
AIS: Acute ischemic stroke
EVT: Endovascular treatment
PSA: Probabilistic sensitivity analysis
ICV: Ischemic core volume
MMR: Core-penumbra mismatch ratio
CTP: CT perfusion
ASPECTS: Alberta Stroke Program Early CT score

## Data sharing statement

The complete de-identified patient datasets from the MR CLEAN-NO IV, and MR CLEAN-MED trials will be available from 18 months after publication until 15 years from publication. Data can be obtained from https://www.contrast-consortium.nl/data-request-form/. The data will be made available to researchers who are CONTRAST consortium members or collaborators, and whose proposed use of the data has been approved by the CONTRAST data access and writing committee. The data will be made available for specified purposes, as defined in the sub study proposal and approved by the CONTRAST data access and writing committee. The data will be made available after approval of the proposal by the CONTRAST data access and writing committee. To ensure publication transparency and quality, researchers should adhere to the CONTRAST publication policy, accessible on https://www.contrast-consortium.nl/publication-policy-contrast/. For the patients included in the MR CLEAN Registry and the local cohort, individual patient data cannot be made available under Dutch law since we did not obtain patient approval for sharing individual patient data. All syntax files and output of statistical analyses are available on reasonable request to the corresponding author.

