## Supplementary material for "Costs and health effects of CT perfusion-based selection for endovascular treatment of patients with a large vessel occlusion presenting within six hours after symptom onset A model-based health economic evaluation": Online Supplement

**Online supplementary material**

**Supplement A: Methods for computing the mRS of a virtual control arm**

To compute the effectiveness of CTP based EVT patient selection a control arm that did not receive EVT is required (noEVT). Since we did not use randomized data, we computed the probability of specific mRS scores of a virtual control arm based on available treatment effect and treatment effect modification odds ratios. Sensitivity analyses were performed for varying values of these odds ratios. For the control arm mRS simulation we need to have the mRS and baseline characteristics an EVT treated patient, a distribution of a population treated with EVT and a control arm (we used the MR CLEAN trial data), and the OR for treatment effect ($OR\left( mRS | EVT \right)=$ 1.67) and treatment effect modification ($OR\left( mRS | EVT*ICV \right)=$ 0.98 per 10 mL) related to mRS. We assume in the example below a patient with mRS 2 after EVT with a core volume at baseline of 60 mL. The following steps were taken:

Based on the ORs we can compute the combined OR for a specific patient that received EVT as if she did not receive EVT (inverse of the OR: formula 1 and 2). Taking the natural log of an OR gives us the absolute mRS shift (formula 3). We can use this absolute shift to compute the effect on outcome of ICV ($ICV(10mL))$ and add this with the effect of noEVT to compute the combined OR (formula 4).

$$OR (mRS|noEVT)=\frac{1}{OR(mRS|EVT)}$$

Formula 1

$$OR (mRS|noEVT*ICV)=\frac{1}{OR(mRS|EVT*ICV)}$$

Formula 2

$$absolute mRS shift=ln(OR)$$

Formula 3

$$OR \left( mRS│noEVT \& ICV \right)=e^{\ln\left( OR \left( mRS | noEVT \right) \right)*1+ln(OR \left( mRS | noEVT*ICV \right)*ICV(per 10mL)}$$

Formula 4

For the next steps we use the $OR(mRS|EVT)$ (=1.67), the total number of patients with EVT in the literature source for treatment effect (MR CLEAN trial n(EVT)=233), the number of patient with EVT achieving mRS≤2 $n(mRS\leq2 \& EVT)$ (=77), the probability of the patients’ mRS given EVT $p(mRS\leq2|EVT)$ (=0.33), the total number of patients in the control arm $n(noEVT)$ (=267), the number of patients in the control arm achieving mRS≤2 $n(mRS\leq2 \& noEVT)$ (=77), and the probability mRS≤2 in the control arm $p(mRS\leq2|noEVT)$ (=0.195). We use the probability and OR for mRS≤2 to get stable estimates irrespective of the simulated patients’ mRS; low sample sizes for mRS 0 and unreasonably large for mRS 6 resulting in inaccurate outcome simulations.

Based on the standard formulas to compute the OR (formulas 5-7) and the new computed combined OR based on literature OR and patient observations from formula 4, it is possible to derive formula 8 for computing $Odds(mRS\leq2|noEVT)$.

$$p(mRS\leq2|EVT)= \frac{n(mRS\leq2 \& EVT)}{n total EVT}$$

Formula 5

$$Odds(mRS\leq2|EVT)= \frac{p(mRS\leq2|EVT)}{1-p(mRS\leq2|EVT)}$$

Formula 6

$$OR(mRS\leq2|EVT)= \frac{Odds(mRS\leq2|EVT)}{Odds(mRS\leq2|noEVT)}$$

Formula 7

$$Odds\left( mRS\leq2 | noEVT \right)= Odds\left( mRS\leq2 | EVT \right)*OR(mRS\leq2|EVT)$$

Formula 8

Finally, after combining formulas 5-8 using a patient specific OR from formula 4 you can compute the virtual probability of mRS shift in the control arm (formula 9 using formulas 5-8). The results per formula for our example are described in the section below formula 9. After solving formula 9, the value of $p\left( mRS\leq2 | noEVT \right)$ is used to compute a 1 point shift. In the example below, if a patient has an mRS of 3 with EVT, if no EVT was given she has a probability of 0.25 for an mRS of 3 and 0.75 for an mRS of 4.

$$p\left( mRS\leq2 | noEVT \right)=(1-p\left( mRS\leq2 | noEVT \right))* Odds\left( mRS\leq2 | noEVT \right)$$

Formula 9

**Example results:**

$$OR \left( mRS | noEVT \right)=\frac{1}{1.67}=0.60$$

Formula 1

$$OR \left( mRS | noEVT*ICV \right)=\frac{1}{0.98}=1.02$$

Formula 2

$$OR \left( mRS│noEVT \& ICV \right)=e^{\ln\left( 0.60 \right)*1+ln(1.02)*6}=0.67$$

Formula 4

$$p\left( mRS\leq2 | EVT \right)= \frac{77}{233}=0.33$$

Formula 5

$$Odds\left( mRS\leq2 | EVT \right)= \frac{0.33}{1-0.33}=0.49$$

Formula 6

$$Odds\left( mRS\leq2 | noEVT \right)= 0.49*0.60=0.33$$

Formula 8

$$p\left( mRS\leq2 | noEVT \right)=(1-p\left( mRS\leq2 | noEVT \right))*0.33$$

Formula 9

$$1.33*p\left( mRS\leq2 | noEVT \right)=0.33$$

$$p\left( mRS\leq2 | noEVT \right)=\frac{0.33}{1.33}=0.25$$

Solving formula 9

The probability of mRS≤2 for the no EVT arm could deviate from observed values in the MR CLEAN trial. This could be due to inaccuracies in the proportional odds assumption or due to different patient characteristics in the EVT arm compared to the no EVT arm.

**Supplement B: Protocol deviations**

The presented work is based on a previously published protocol paper^1^ and the corresponding protocol online (trial registration number: NL7974; trialsearch.who.int/Trial2.aspx?TrialID=NL7974). Below are the deviations to the protocol described with motivation.

**Default odds ratio EVT effect**

In the protocol, we mistakenly reported a default scenario OR of 1.86 (95%CI:1.34-2.59) for EVT effect. This should have been 1.67 (95%CI:1.21-2.3) as reported as adjusted common OR in the MR CLEAN trial. Since a lower EVT effect is beneficial for the use of CTP for EVT patient selection (and findings were negative regardless) we used the OR of 1.67. In additional sensitivity analyses (table 1), the value of 1.97 was also considered.

**Probabilistic simulations**

We reduced the number of resamples from 10,000 to 1,000 for computational purposes. As the proportion of patients excluded for EVT was relatively low due to the low occurrence of unfavorable perfusion profiles, we altered the sampled cohort size of our initial simulation to the original cohort size (n=703) instead of 100.

**Analyses**

We included analyses for the core-penumbra mismatch ratio up to present a more complete overview of potential CTP uses. Furthermore, we added a 10-year follow-up horizon and a most favorable CTP scenario considering elderly (aged≥80 years) with a low EVT effect and high effect modification due to CTP parameters. We found that these analyses were of added value as they are sometimes considered and, even in the extreme cases, did not alter the conclusion.

**Primary outcome measure**

Excluding patients for EVT resulted at best in a cost savings for a limited loss of health in a small group of patients in the simulated cohorts. Therefore, ΔQALYs per patient as average over the simulated cohorts was often zero or close to zero resulting in an ICER (ΔCosts/ ΔQALYs) going to infinity or impossible values (it is impossible to divide by zero). Thus, we chose to use the NMB as primary outcome measure as this metric is insensitive to zero or close to zero values for ΔQALYs. In appendix G, the ICER for all simulations is also reported together with the ΔCosts and ΔQALYs.

**Supplement C: CTP acquisition, post-processing, and quality assessment**

CTP scans were acquired according to local acquisition protocols, which differed per study site. CTP was collected from the same stroke centers for all sources in CLEOPATRA. Perfusion analysis was centrally performed by a trained observer using syngo.via CT Neuro Perfusion (version VB40, Siemens Healthcare, Forchheim, Germany). The ICV was estimated using a threshold CBV <1.2 mL/100 mL and the critically hypoperfused tissue (including penumbra) was defined as CBF <27 mL/100 mL/min. A default smoothing filter was applied (smoothing strength 10 mm)^2^. These approaches were chosen based on previous research which has shown that these settings have the best agreement with RAPID software, which is a commonly used perfusion analysis software also used for the HERMES analysis on CTP perfusion ^2,3^. Visual quality assessment of the CTP results was performed by two experienced neuroradiologists (>10 and >15 years of experience). In patients with an estimated CTP ischemic core volume ≥70 mL, craniocaudal cropping of the core segmentation was performed by an experienced reader to remove obvious artifacts at the level of the skull base. Quality of the CTP data (i.e. Z-axis coverage of the brain, adequate tube voltage, adequate contrast bolus injection) was comparable between all data sources in CLEOPATRA. The penumbra (or mismatch) volume was defined as critically hypoperfused volume minus ischemic core volume. The core-penumbra mismatch ratio was calculated as the critically hypoperfused volume divided by the ischemic core volume.

**Supplement D: Model input variables**

The table below describes all the input variables used for modelling purposes. This table contains alterations from our previous publication.^1^

| **Variable** | **Value** | **Distribution** | **Data source** |
| --- | --- | --- | --- |
| EVT effect (odds ratio) | 1.67 (95%CI:1.21-2.3) (baseline)  1.97 (95%CI:1.51-2.6) (upper)  1.37 (95%CI:0.91-2.0) (lower)  1.17 (95%CI:0.71-1.8) (very low)  2.49 (95%CI:1.76-3.53) (HERMES) | Log-normal | Berkhemer et al.^4^  Goyal et al.^5^ |
| EVT effect modification of core volume (ICV*EVT odds ratio) | 0.98 (95%CI:0.881-1.091) (baseline)  0.93 (95%CI:0.831-1.041)  0.88 (95%CI:0.781-0.991) | Log-normal | New analysis HERMES data^5^, Table D.2 |
| EVT effect modification of core-penumbra mismatch ratio (MMR*EVT odds ratio) | 1.010 (95%CI:0.994;1.560) (baseline)  1.060 (95%CI:1.044;1.610)  1.110 (95%CI:1.094;1.660) | Log-normal | New analysis HERMES data^5^, Table D.3 |
| CTP ischemic core volume treatment decision threshold | Varying between 0-150 per 10 mL  70 mL at baseline | Fixed value | Deterministic |
| CTP core-penumbra mismatch ratio treatment decision threshold | Varying between 1.2-2.4 per 0.2  1.8 at baseline | Fixed value | Deterministic |
| Baseline probability of stroke recurrence | Dependent on years after index  ischemic stroke | Fixed value | Pennlert et al.^6^ |
| HR recurrent stroke | age and mRS dependent | Log-normal | Pennlert et al.^6^ |
| Baseline probability of death | age, gender and year dependent | Fixed value | Dutch Royal Actuarian Society^7^ |
| HR mortality (by mRS: 01/2/3/4/5) | 1.54/2.17/3.18/4.55/6.55 | Log-normal | Hong et al.^8^ |
| Inflation rate in % per year | % per year  Assumed for 2022: 9% | Fixed value | Central Bureau of Statistics^9^ |
| Costs EVT* | €9924.50 | Fixed value | Van den Berg et al.^10^ |
| Costs IVT* | €950.82 | Fixed value | Van den Berg et al.^10^ |
| Costs of CTP* | €251.40 | Fixed value | Estimated in protocol^1^ |
| Costs year 1 (by 90 day mRS: 01/2/3/4/5/6)* | 33,402(31,930)/52,804(23,571)/82,452(35,333)/  112,414(35,786)/96,640(30,463)/21,112(17,350) | Gamma | Van Voorst et al.^11^ |
| Costs year 2 (by 18 month mRS: 01/2/3/4/5/6)* | 5,934(15,918)/8,543(14,844)/19,235(15,999)/  43,193(45,640)/56,425)/24,252)/423(3,196) | Gamma | Van Voorst et al.^11^ |
| Costs year 2 onward (by 18 month mRS: 01/2/3/4/5/6)* | 3,633(9,087)/7,318(13,770)/16,276(11,753)/  31,037(19,928)/54,997(24,874)/374(3,118) | Gamma | Van Voorst et al.^11^ |
| QALY (by mRS: 01/2/3/4/5/6) | 0.94(0.09)/0.80(0.17)/0.68(0.24)/  0.39(0.26)/0.24(0.25)/0(0.01) | Beta | Van Voorst et al.^11^ |

**Table D.1. Data sources for model input parameter estimation*.** EVT: endovascular treatment; HR: hazard ratio; IVT: intravenous thrombolysis; mRS: modified Rankin Scale score; OR: odds ratio. *Costs are depicted for the reference year 2015.

**Cost data specification**

Data described below has previously been published.^1,10,11^

*Costs per unit of care:* The Dutch costing manual for health care research^12^*,* hospital registration systems, and data published by the institute of Medical Technological Assessment (iMTA, Rotterdam, the Netherlands) were used to extract costs per unit of care for the reference year 2015. The total costs of acute care and follow-up healthcare costs were computed based on unit of care volume estimates. All acute costs were inflated by 42% to account for overhead costs of the hospital (iMTA guideline).

*Acute care units of care*:

*Costs for CTP* (€251.50) were estimated in the protocol paper: 10% additional costs for acute care personnel (€94), €129 for the CTP acquisition,^12^ and €20 for the CTP software license costs per patient. The total was inflated by 42% to accure for overhead costs.

*Costs for endovascular treatment* (€9924.50): Costs for 1 CTA and NCCT scan were added. Personal costs included 1 (neuro-) interventionist, 1 anesthesiologist, 2 radiology assistants, 2 anesthesia assistants during 1.5 hours of treatment delivery. We assumed that all procedures were conducted with general anesthesia as this added only marginal costs. Material during the endovascular procedure was assumed to consist of thrombectomy materials (guide wire, balloon, stent retriever or aspiration device), angiographic materials, vascular closure devices, and heparin.

*Costs for intravenous thrombolysis* (€950.82) were added if patients received it. The costs of thrombolysis delivery were extracted from Medicijnkosten.nl 2016.

*Long-term costs*

The following cost of care were considered: In-hospital care use, outpatient clinic visits, rehabilitation, formal

homecare, and long-term institutionalized care. The units of these long-term posts were estimated with patient questionnaires and report forms, medical records, and the hospital information systems.^10^ We computed the costs per mRS at 3 months for the first year and at 18 months for the second year. The long-term costs of the third year onward were assumed the same as the costs for the second year without rehabilitation costs.

**Quality adjusted life-years (QALYs)**

QALYs were computed per mRS sub-score per year based on 391 patients with 2-year follow-up.^10^ EuroQol5D (EQ5D) questionnaires were used to estimate the QALYs at 3, 6, 9, 12, 18, and 24 months after acute ischemic stroke. The resulting value was used as the QALYs per unit of time in an mRS group was computed per patient by averaging the EQ%D utility across different time points per mRS group

| **Effect** | **OR** | **Lower bound 95%CI** | **Upper bound 95%CI** | **p-value** |
| --- | --- | --- | --- | --- |
| Age (per 5 yr) | 0.834 | 0.786 | 0.885 | <0.001 |
| NIHSS (per 5 pts) | 0.815 | 0.699 | 0.949 | 0.009 |
| Onset to randomization  (per 30 min) | 0.950 | 0.901 | 1.002 | 0.059 |
| ASPECTS | 1.077 | 0.979 | 1.185 | 0.127 |
| Female gender | 0.812 | 0.600 | 1.098 | 0.175 |
| tPA administered | 0.923 | 1.508 | 0.565 | 0.748 |
| ICA (vs M2) | 0.468 | 0.226 | 0.969 | 0.041 |
| M1 (vs M2) | 0.709 | 0.364 | 1.379 | 0.310 |
| EVT (vs best medical management) | 2.520 | 1.483 | 4.283 | 0.001 |
| Ischaemic core volume  (per 10 ml) | 0.862 | 0.795 | 0.935 | <0.001 |
| EVT * Ischaemic core volume  (per 10 ml) | 0.980 | 0.881 | 1.091 | 0.717 |

**Table D.2. Endovascular treatment effect modification for ordinal function outcome due to ischaemic core volume.** Presented results were extracted in a patient level analysis of the HERMES pooling. This analysis was similar to the one described by Campbell et al. ^3^. 95%CI: 95% confidence interval. ASPECTS: Alberta stroke program early CT score, NIHSS: National institute of health stroke. ICA: internal carotid artery. M1/M2: segments of middle cerebral artery. IVT: intravenous thrombolysis. EVT: endovascular treatment. MMR: core-penumbra mismatch ratio.

| **Effect** | **OR** | **Lower bound 95%CI** | **Upper bound 95%CI** | **p-value** |
| --- | --- | --- | --- | --- |
| Age (per 5 yr) | 0.818 | 0.763 | 0.876 | <0.0001 |
| NIHSS (per 5 pts) | 0.702 | 0.591 | 0.833 | <0.0001 |
| Onset to randomization  (per 30 min) | 0.959 | 0.905 | 1.017 | 0.164 |
| ASPECTS | 1.106 | 1.000 | 1.223 | 0.050 |
| Female gender | 0.967 | 0.695 | 1.346 | 0.844 |
| IVT | 0.994 | 0.584 | 1.693 | 0.983 |
| ICA (vs M2) | 0.385 | 0.172 | 0.861 | 0.020 |
| M1 (vs M2) | 0.553 | 0.264 | 1.159 | 0.116 |
| EVT (vs best medical management) | 1.984 | 1.055 | 3.730 | 0.033 |
| MMR (per 1 point) | 1.003 | 0.990 | 1.015 | 0.677 |
| EVT * MMR (per 1 point) | 1.010 | 0.994 | 1.026 | 0.228 |

**Table D.3. Endovascular treatment effect modification for ordinal function outcome due to mismatch ratio.** Presented results were extracted in a patient level analysis of the HERMES pooling. This analysis was similar to the one described by Campbell et al. ^3^. Only patients with an ischaemic core volume > 0 were considered. 95%CI: 95% confidence interval. ASPECTS: Alberta stroke program early CT score, NIHSS: National institute of health stroke. ICA: internal carotid artery. M1/M2: segments of middle cerebral artery. IVT: intravenous thrombolysis. EVT: endovascular treatment. MMR: core-penumbra mismatch ratio.

**Supplement E: Inclusion and baseline characteristics**

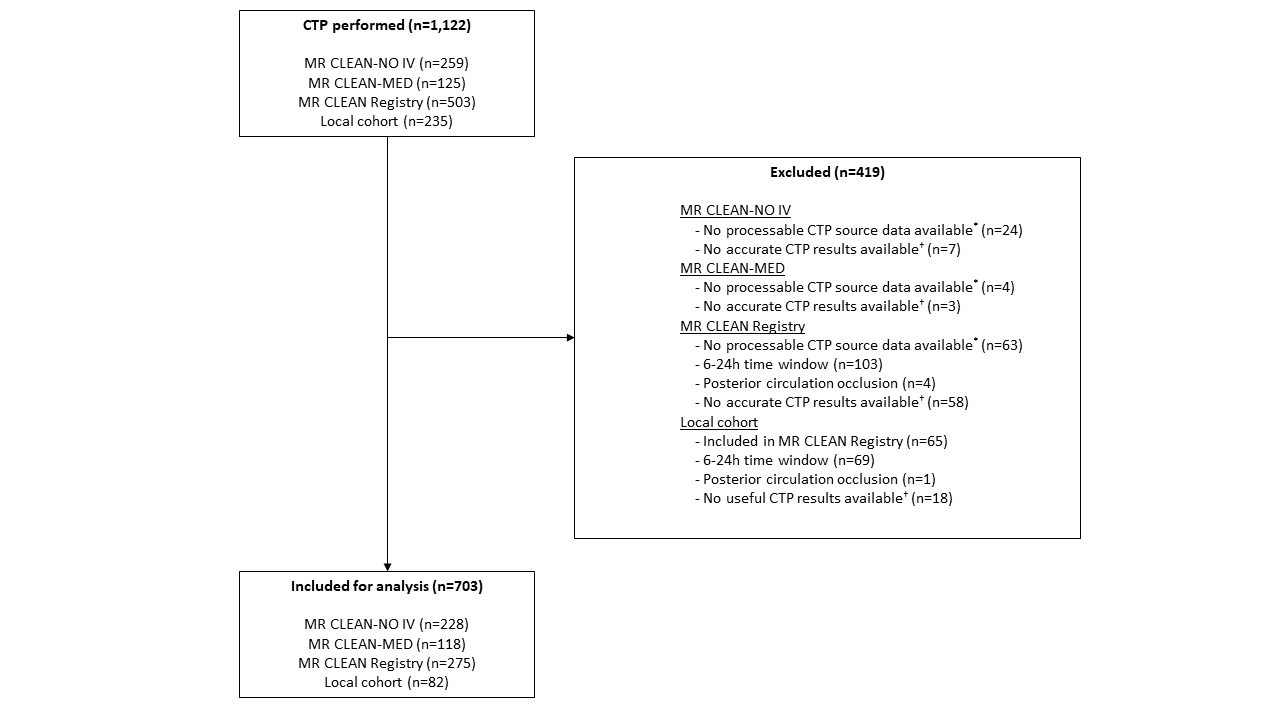

**Figure E1. Flowchart of patient selection.** CTP: CT perfusion. ^*^CTP source data without time information or CTP source data not available due to local storage in primary stroke centre. ^†^Reasons for inaccurate CTP results include severe patient motion, severe curve truncation, no timely contrast arrival or incorrect timing CTP, or severe artefacts in CTP source data.

| **Variables** | **Values** |
| --- | --- |
| **Age**  **median (IQR)** | 72 (62;81) |
| **Male sex**  **number (%)** | 391 (55·6%) |
| **Baseline NIHSS**  **median (IQR)** | 15 (9;19) |
| **mRS before AIS**  **number (%)** |  |
| **0** | 404 (64.4%) |
| **1** | 116 (18.5%) |
| **2** | 63 (10.0%) |
| **≥3** | 44 (7.0%) |
| **Baseline ASPECTS**  **median (IQR)** | 9 (8;10) |
| **Baseline collateral score**  **number (%)** |  |
| **0** | 30 (4.8%) |
| **1** | 191 (30.9%) |
| **2** | 281 (45.4%) |
| **3** | 117 (18.9%) |
| **Occlusion location**  **number (%)** |  |
| **ICA** | 41 (5·8%) |
| **ICA-T** | 127 (18·1%) |
| **M1** | 367 (52·4%) |
| **M2** | 166 (23.7%) |
| **Ischaemic core volume in mL**  **median (IQR)** | 13 (5;33) |
| **Penumbra volume in mL**  **Mean (SD)** | 113 (128) |
| **Core-penumbra mismatch ratio**  **median (IQR)** | 9 (4;19) |
| **Onset-to-groin time in minutes**  **median (IQR)** | 158 (115;230) |
| **Duration of procedure in minutes**  **median (IQR)** | 48 (31;71) |
| **Received intravenous treatment with alteplase (0.9mg/kg)**  **number (%)** | 382 (67.0%) |
| **mRS 90-days after EVT**  **number (%)** |  |
| **0** | 47 (7.3%) |
| **1** | 116 (17.9%) |
| **2** | 154 (23.8%) |
| **3** | 70 (10.8%) |
| **4** | 76 (11.7%) |
| **5** | 42 (6.5%) |
| **6** | 142 (21.9%) |

**Table E1: Baseline characteristics of the included cohort.** Abbreviations: IQR: interquartile range, SD: standard deviation, ASPECTS: Alberta stroke programme early CT score, NIHSS: National institute of health stroke, mRS: modified Rankin Scale, AIS: acute ischemic stroke, eTICI: extended Thrombolysis in Cerebral Ischemia.

| **Variable** | **Ischemic core volume (mL) subgroup** | | | | | | | | | **p-value** |
| --- | --- | --- | --- | --- | --- | --- | --- | --- | --- | --- |
|  | **≤10** | **10-20** | **20-30** | **30-40** | **40-50** | **50-70** | **70-90** | **90-110** | **≥110** |  |
| **Number of patients** | 301 | 134 | 78 | 54 | 30 | 35 | 33 | 14 | 23 |  |
| **Age (median - IQR)** | 71 (61;80) | 73 (65;83) | 69 (64;81) | 68 (56;80) | 72 (60;76) | 74 (64;82) | 65 (57;76) | 66 (48;74) | 73 (68;79) | 0.13 |
| **Sex** |  |  |  |  |  |  |  |  |  | <1e-5 |
| **Female** | 169 (56.5%) | 59 (44.0%) | 24 (31.2%) | 14 (25.9%) | 9 (30.0%) | 13 (37.1%) | 11 (33.3%) | 5 (35.7%) | 6 (26.1%) |  |
| **Male** | 130 (43.5%) | 75 (56.0%) | 53 (68.8%) | 40 (74.1%) | 21 (70.0%) | 22 (62.9%) | 22 (66.7%) | 9 (64.3%) | 17 (73.9%) |  |
| **Baseline NIHSS (median - IQR)** | 12 (7;17) | 15 (10;19) | 16 (11;19) | 17 (13;20) | 18 (14;22) | 20 (16;22) | 20 (17;23) | 17 (16;21) | 19 (17;23) | <1e-5 |
| **mRS prior to AIS** |  |  |  |  |  |  |  |  |  | 0.92 |
| **0** | 161 (60.8%) | 81 (66.4%) | 49 (73.1%) | 29 (61.7%) | 19 (65.5%) | 19 (61.3%) | 23 (74.2%) | 8 (66.7%) | 15 (68.2%) |  |
| **1** | 57 (21.5%) | 22 (18.0%) | 9 (13.4%) | 8 (17.0%) | 5 (17.2%) | 5 (16.1%) | 4 (12.9%) | 2 (16.7%) | 3 (13.6%) |  |
| **2** | 31 (11.7%) | 9 (7.4%) | 6 (9.0%) | 7 (14.9%) | 3 (10.3%) | 4 (12.9%) | 2 (6.5%) | 0 (0.0%) | 1 (4.5%) |  |
| **≥3** | 16 (6.0%) | 10 (8.2%) | 3 (4.5%) | 3 (6.4%) | 2 (6.9%) | 3 (9.7%) | 2 (6.5%) | 2 (16.7%) | 3 (13.6%) |  |
| **Baseline ASPECTS (median-IQR)** | 10 (9;10) | 9 (8;10) | 9 (7;10) | 9 (7;10) | 9 (8;9) | 8 (6;10) | 9 (7;10) | 9 (7;9) | 8 (5;10) | <1e-5 |
| **Baseline collateral score** |  |  |  |  |  |  |  |  |  | <1e-5 |
| **0** | 1 (0.4%) | 3 (2.5%) | 4 (6.0%) | 3 (6.5%) | 0 (0.0%) | 4 (12.9%) | 4 (12.5%) | 2 (15.4%) | 9 (40.9%) |  |
| **1** | 47 (17.9%) | 31 (26.1%) | 23 (34.3%) | 21 (45.7%) | 14 (53.8%) | 16 (51.6%) | 21 (65.6%) | 9 (69.2%) | 9 (40.9%) |  |
| **2** | 145 (55.3%) | 60 (50.4%) | 30 (44.8%) | 18 (39.1%) | 8 (30.8%) | 8 (25.8%) | 7 (21.9%) | 2 (15.4%) | 2 (9.1%) |  |
| **3** | 69 (26.3%) | 25 (21.0%) | 10 (14.9%) | 4 (8.7%) | 4 (15.4%) | 3 (9.7%) | 0 (0.0%) | 0 (0.0%) | 2 (9.1%) |  |
| **Occlusion location** |  |  |  |  |  |  |  |  |  | <0.001 |
| **ICA** | 19 (6.3%) | 11 (8.2%) | 4 (5.2%) | 1 (1.9%) | 2 (6.7%) | 0 (0.0%) | 1 (3.0%) | 1 (7.1%) | 2 (8.7%) |  |
| **ICA-T** | 41 (13.7%) | 16 (11.9%) | 17 (22.1%) | 10 (18.5%) | 4 (13.3%) | 14 (40.0%) | 10 (30.3%) | 6 (42.9%) | 9 (39.1%) |  |
| **M1** | 169 (56.3%) | 73 (54.5%) | 31 (40.3%) | 32 (59.3%) | 13 (43.3%) | 16 (45.7%) | 20 (60.6%) | 4 (28.6%) | 9 (39.1%) |  |
| **M2** | 71 (23.7%) | 34 (25.4%) | 25 (32.5%) | 11 (20.4%) | 11 (36.7%) | 5 (14.3%) | 2 (6.1%) | 3 (21.4%) | 3 (13.0%) |  |
| **Ischemic core volume in mL (median – IQR)** | 4 (2;7) | 14 (12;17) | 24 (22;27) | 35 (33;38) | 45 (43;48) | 61 (57;66) | 77 (72;82) | 94 (92;100) | 154 (123;196) | <1e-5 |
| **Penumbra volume in mL (mean - SD)** | 107 (188) | 110 (51) | 119 (48) | 116 (45) | 132 (45) | 124 (55) | 126 (43) | 131 (44) | 108 (53) | 0.95 |
| **Core-penumbra mismatch ratio (median - IQR)** | 19 (13;36) | 9 (6;12) | 6 (5;8) | 4 (3;5) | 4 (3;5) | 3 (2;4) | 3 (2;4) | 2 (2;2) | 2 (2;2) | <0.001 |
| **Onset to groin time in minutes (median - IQR)** | 155 (115;225) | 162 (122;254) | 145 (115;209) | 158 (111;212) | 158 (117;226) | 165 (110;194) | 192 (125;307) | 163 (120;195) | 166 (118;212) | 0.83 |
| **Duration of procedure in minutes (median - IQR)** | 49 (31;75) | 40 (30;66) | 42 (29;62) | 56 (28;83) | 47 (32;61) | 55 (39;80) | 50 (35;64) | 55 (40;85) | 56 (40;66) | 0.60 |
| **Intravenous thrombolysis administration** |  |  |  |  |  |  |  |  |  | 0.44 |
| **No** | 69 (28.5%) | 40 (36.7%) | 22 (33.8%) | 12 (27.3%) | 11 (42.3%) | 13 (48.1%) | 11 (39.3%) | 4 (36.4%) | 6 (33.3%) |  |
| **Yes** | 173 (71.5%) | 69 (63.3%) | 43 (66.2%) | 32 (72.7%) | 15 (57.7%) | 14 (51.9%) | 17 (60.7%) | 7 (63.6%) | 12 (66.7%) |  |
| **mRS 90-days after EVT** |  |  |  |  |  |  |  |  |  | <0.001 |
| **0** | 30 (10.7%) | 9 (7.3%) | 2 (2.9%) | 3 (6.4%) | 1 (3.8%) | 0 (0.0%) | 2 (6.1%) | 0 (0.0%) | 0 (0.0%) |  |
| **1** | 58 (20.7%) | 26 (21.1%) | 16 (23.2%) | 8 (17.0%) | 3 (11.5%) | 1 (3.1%) | 4 (12.1%) | 0 (0.0%) | 0 (0.0%) |  |
| **2** | 78 (27.9%) | 30 (24.4%) | 16 (23.2%) | 11 (23.4%) | 4 (15.4%) | 4 (12.5%) | 8 (24.2%) | 1 (7.1%) | 2 (9.1%) |  |
| **3** | 26 (9.3%) | 14 (11.4%) | 9 (13.0%) | 4 (8.5%) | 5 (19.2%) | 3 (9.4%) | 5 (15.2%) | 3 (21.4%) | 1 (4.5%) |  |
| **4** | 23 (8.2%) | 19 (15.4%) | 7 (10.1%) | 8 (17.0%) | 4 (15.4%) | 4 (12.5%) | 7 (21.2%) | 3 (21.4%) | 1 (4.5%) |  |
| **5** | 21 (7.5%) | 4 (3.3%) | 3 (4.3%) | 2 (4.3%) | 2 (7.7%) | 3 (9.4%) | 3 (9.1%) | 1 (7.1%) | 3 (13.6%) |  |
| **6** | 44 (15.7%) | 21 (17.1%) | 16 (23.2%) | 11 (23.4%) | 7 (26.9%) | 17 (53.1%) | 4 (12.1%) | 6 (42.9%) | 15 (68.2%) |  |

**Table E2: Baseline characteristics of the CLEOPATRA population per 10 mL ischemic core volume.** Abbreviations: IQR: interquartile range, SD: standard deviation, ASPECTS: Alberta stroke programme early CT score, NIHSS: National institute of health stroke, mRS: modified Rankin Scale, AIS: acute ischemic stroke, eTICI: extended Thrombolysis in Cerebral Ischemia.

| **Variable** | **Core-penumbra mismatch ratio (penumbra volume/core volume)** | | | | | | |  |
| --- | --- | --- | --- | --- | --- | --- | --- | --- |
|  | **<=1.4** | **1.4-1.6** | **1.6-1.8** | **1.8-2.0** | **2.0-2.2** | **2.2-2.4** | **>=2.4** | **p-value** |
| **Number of patients** | 5 | 6 | 8 | 11 | 15 | 19 | 635 |  |
| **Age (median - IQR)** | 80 (71;81) | 72 (69;75) | 69 (50;79) | 73 (58;78) | 64 (63;82) | 68 (58;73) | 72 (62;81) | 0.66 |
| **Sex** |  |  |  |  |  |  |  | 0.32 |
| **Female** | 3 (60.0%) | 1 (16.7%) | 3 (37.5%) | 2 (18.2%) | 7 (46.7%) | 6 (31.6%) | 287 (45.2%) |  |
| **Male** | 2 (40.0%) | 5 (83.3%) | 5 (62.5%) | 9 (81.8%) | 8 (53.3%) | 13 (68.4%) | 348 (54.8%) |  |
| **Baseline NIHSS (median - IQR)** | 18 (15;25) | 18 (18;22) | 20 (16;23) | 18 (16;20) | 17 (16;20) | 18 (13;20) | 15 (9;19) | <0.01 |
| **mRS prior to AIS** |  |  |  |  |  |  |  | 0.85 |
| **0** | 2 (50.0%) | 4 (66.7%) | 6 (75.0%) | 8 (80.0%) | 7 (58.3%) | 11 (64.7%) | 364 (64.3%) |  |
| **1** | 1 (25.0%) | 1 (16.7%) | 1 (12.5%) | 1 (10.0%) | 1 (8.3%) | 2 (11.8%) | 107 (18.9%) |  |
| **2** | 1 (25.0%) | 1 (16.7%) | 0 (0.0%) | 0 (0.0%) | 2 (16.7%) | 1 (5.9%) | 58 (10.2%) |  |
| **≥3** | 0 (0.0%) | 0 (0.0%) | 1 (12.5%) | 1 (10.0%) | 2 (16.7%) | 3 (17.6%) | 37 (6.5%) |  |
| **Baseline ASPECTS (median-IQR)** | 8 (5;8) | 10 (7;10) | 8 (8;9) | 10 (7;10) | 10 (8;10) | 8 (7;10) | 9 (8;10) | 0.07 |
| **Baseline collateral score** |  |  |  |  |  |  |  | <1e-5 |
| **0** | 4 (80.0%) | 3 (60.0%) | 2 (25.0%) | 1 (12.5%) | 0 (0.0%) | 1 (5.3%) | 19 (3.4%) |  |
| **1** | 1 (20.0%) | 2 (40.0%) | 3 (37.5%) | 5 (62.5%) | 6 (42.9%) | 12 (63.2%) | 161 (29.0%) |  |
| **2** | 0 (0.0%) | 0 (0.0%) | 2 (25.0%) | 2 (25.0%) | 6 (42.9%) | 5 (26.3%) | 265 (47.7%) |  |
| **3** | 0 (0.0%) | 0 (0.0%) | 1 (12.5%) | 0 (0.0%) | 2 (14.3%) | 1 (5.3%) | 111 (20.0%) |  |
| **Occlusion location** |  |  |  |  |  |  |  | 0.12 |
| **ICA** | 0 (0.0%) | 1 (16.7%) | 0 (0.0%) | 1 (9.1%) | 2 (13.3%) | 0 (0.0%) | 37 (5.8%) |  |
| **ICA-T** | 4 (80.0%) | 1 (16.7%) | 2 (25.0%) | 3 (27.3%) | 4 (26.7%) | 6 (31.6%) | 107 (16.9%) |  |
| **M1** | 1 (20.0%) | 3 (50.0%) | 4 (50.0%) | 3 (27.3%) | 5 (33.3%) | 8 (42.1%) | 342 (54.0%) |  |
| **M2** | 0 (0.0%) | 1 (16.7%) | 2 (25.0%) | 4 (36.4%) | 4 (26.7%) | 5 (26.3%) | 147 (23.2%) |  |
| **Ischemic core volume in mL (median – IQR)** | 208 (184;246) | 157 (132;200) | 109 (94;121) | 43 (18;70) | 64 (45;88) | 70 (39;87) | 12 (5;26) | <1e-5 |
| **Penumbra volume in mL (mean - SD)** | 78 (25) | 89 (33) | 76 (23) | 71 (44) | 86 (31) | 82 (35) | 116 (134) | 0.60 |
| **Onset to groin time in minutes (median - IQR)** | 225 (145;268) | 166 (116;197) | 158 (118;194) | 135 (120;172) | 182 (146;239) | 177 (145;272) | 155 (115;230) | 0.72 |
| **Duration of procedure in minutes (median - IQR)** | 60 (30;66) | 52 (40;60) | 55 (43;89) | 56 (42;72) | 54 (30;62) | 62 (38;90) | 47 (30;71) | 0.57 |
| **Intravenous thrombolysis administration** |  |  |  |  |  |  |  | 0.12 |
| **No** | 3 (75.0%) | 0 (0.0%) | 1 (14.3%) | 1 (14.3%) | 5 (35.7%) | 9 (52.9%) | 168 (32.6%) |  |
| **Yes** | 1 (25.0%) | 3 (100.0%) | 6 (85.7%) | 6 (85.7%) | 9 (64.3%) | 8 (47.1%) | 347 (67.4%) |  |
| **mRS 90-days after EVT** |  |  |  |  |  |  |  | <0.0001 |
| **0** | 0 (0.0%) | 0 (0.0%) | 0 (0.0%) | 1 (9.1%) | 0 (0.0%) | 0 (0.0%) | 46 (7.9%) |  |
| **1** | 0 (0.0%) | 0 (0.0%) | 0 (0.0%) | 3 (27.3%) | 0 (0.0%) | 1 (5.9%) | 112 (19.2%) |  |
| **2** | 0 (0.0%) | 1 (16.7%) | 0 (0.0%) | 1 (9.1%) | 2 (14.3%) | 2 (11.8%) | 146 (25.1%) |  |
| **3** | 1 (20.0%) | 0 (0.0%) | 2 (25.0%) | 0 (0.0%) | 1 (7.1%) | 2 (11.8%) | 64 (11.0%) |  |
| **4** | 0 (0.0%) | 0 (0.0%) | 1 (12.5%) | 0 (0.0%) | 4 (28.6%) | 4 (23.5%) | 67 (11.5%) |  |
| **5** | 0 (0.0%) | 0 (0.0%) | 2 (25.0%) | 1 (9.1%) | 0 (0.0%) | 3 (17.6%) | 36 (6.2%) |  |
| **6** | 4 (80.0%) | 5 (83.3%) | 3 (37.5%) | 5 (45.5%) | 7 (50.0%) | 5 (29.4%) | 111 (19.1%) |  |

**Table E3: Baseline characteristics of the CLEOPATRA population by core-penumbra mismatch ratio.** For 4 patients to mismatch ratio values were available. Abbreviations: IQR: interquartile range, SD: standard deviation, ASPECTS: Alberta stroke program early CT score, NIHSS: National institute of health stroke, mRS: modified Rankin Scale, AIS: acute ischemic stroke, eTICI: extended Thrombolysis in Cerebral Ischemia.

**Supplement F: Extended result figures for CTP based EVT selection**

Extended sensitivity analyses considering extended horizon simulations (10 year), per decision threshold the outcome measures, extended measures for the per OR increase or decrease of EVT effect and ICV or MMR based effect modification.

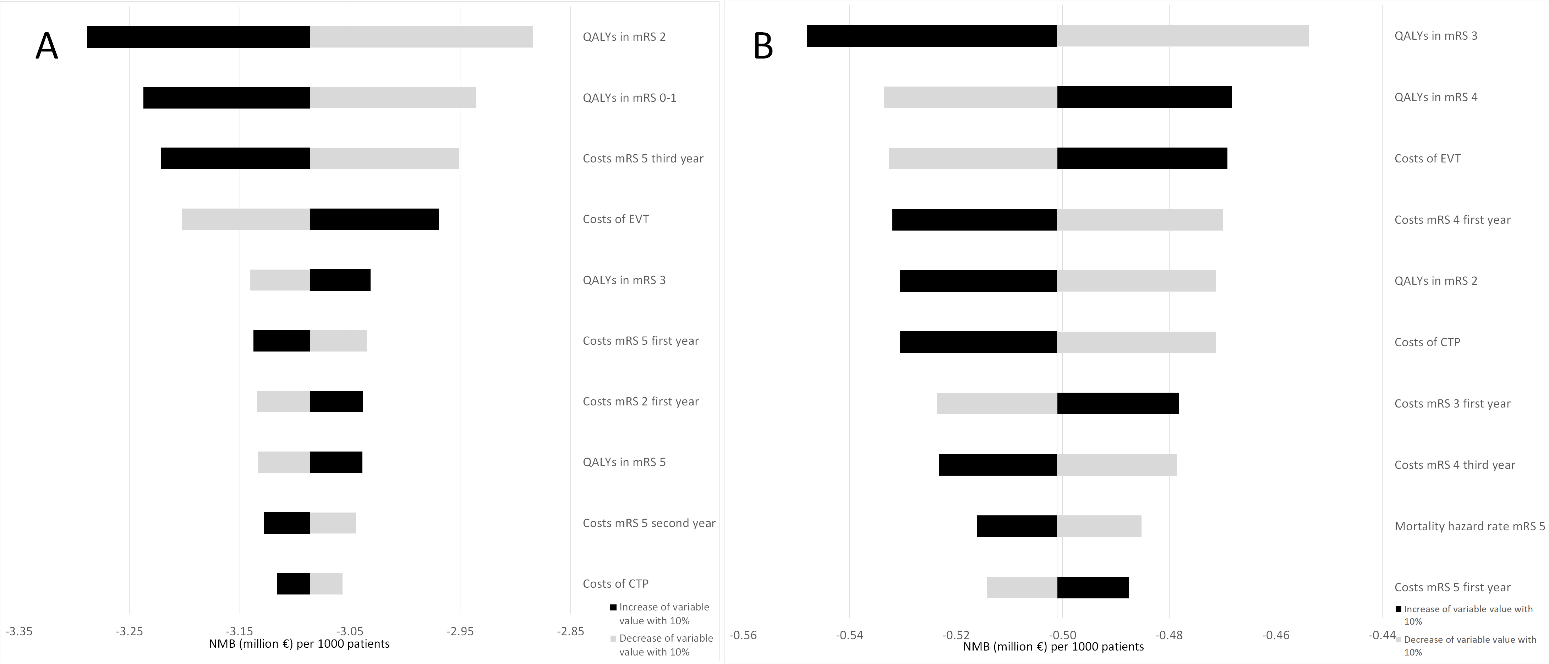

**Figure F.1: Tornado diagram of the one-way sensitivity analyses.** Changes in NMB due to a 10% increase (black) and decrease (gray) relative to the baseline simulation for the 10 most influential model input variables. The input ORs for EVT effect, ICV-, and MMR-based EVT effect modification were not considered in the one-way sensitivity analyses. A 5-year horizon was used. 3A: One-way sensitivity analysis using a baseline ICV threshold of ≥70mL to exclude patients for EVT. 3B: One-way sensitivity analysis using a baseline MMR threshold of ≤1.8 to exclude patients for EVT. NMB: net monetary benefit at a willingness to pay of €80,000 per QALY. ICV: ischemic core volume. EVT: endovascular treatment. QALY: quality-adjusted life-years. mRS: modified Rankin Scale. CTP: CT perfusion. EVT: endovascular treatment.

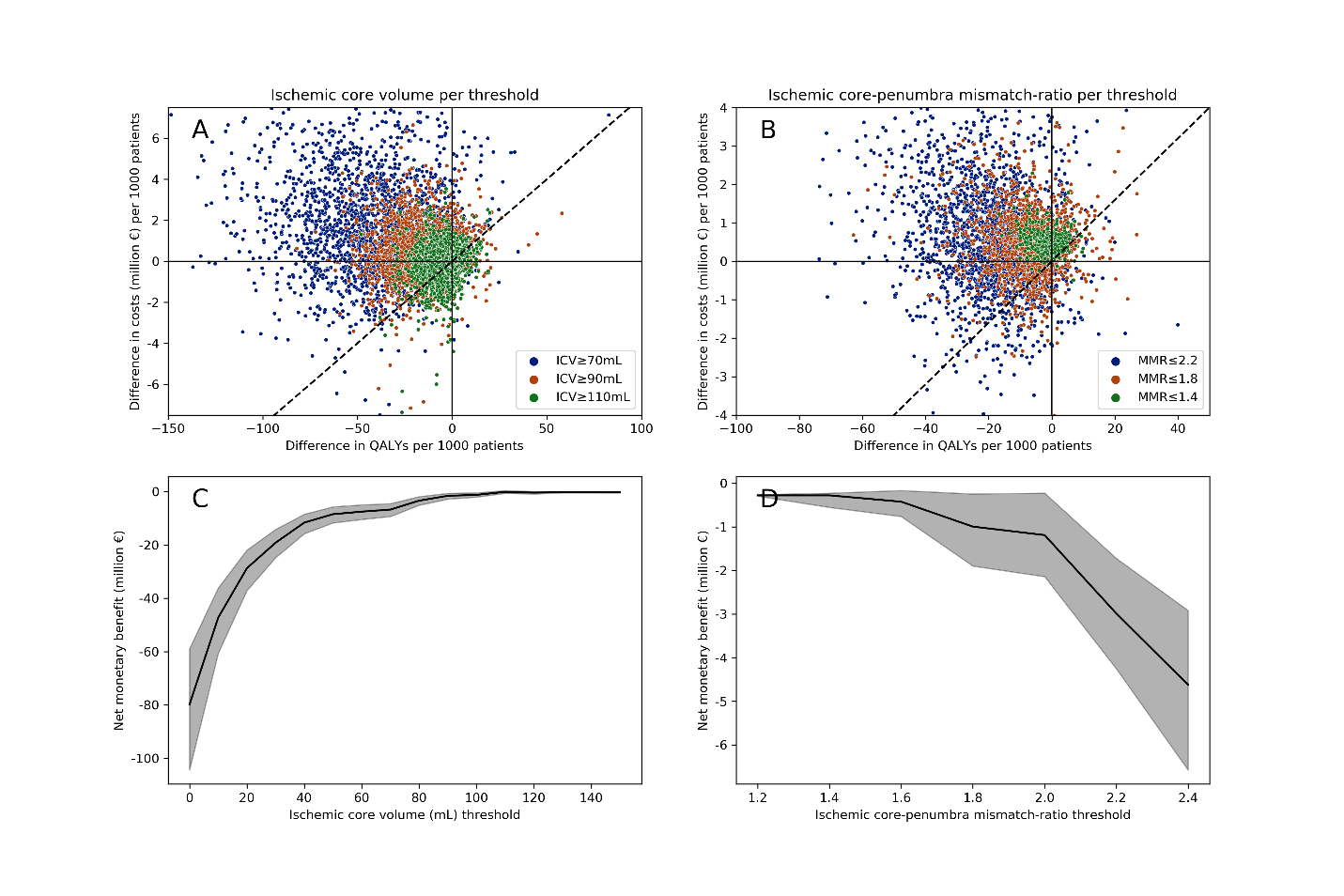

**Figure F.2: 10-year follow-up horizon baseline PSA results per 1000 patients.** This figure is similar to figure 4 in the main article but with 10-year follow-up.

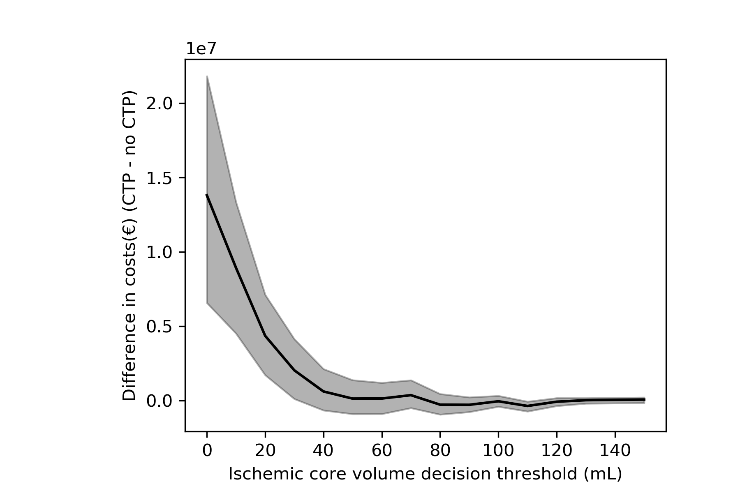

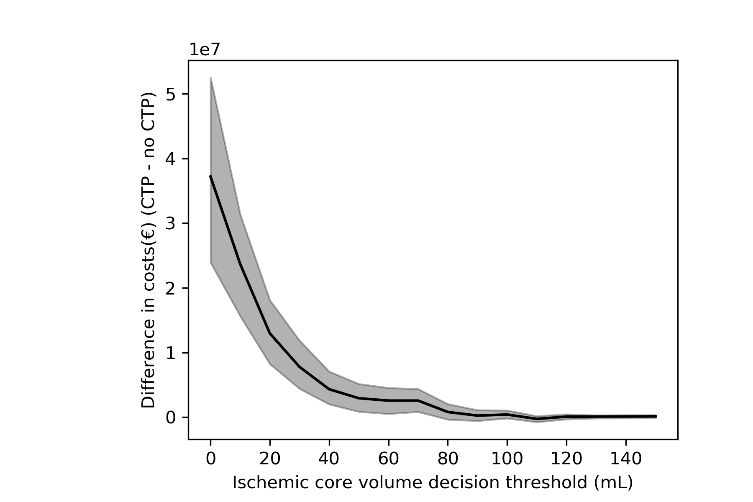

**Figure F.3: ΔCosts per ischemic core volume threshold per 1000 patients.** Values are presented as median with IQR for the baseline PSA. Left: 5-year follow-up. Right: 10 year follow-up.

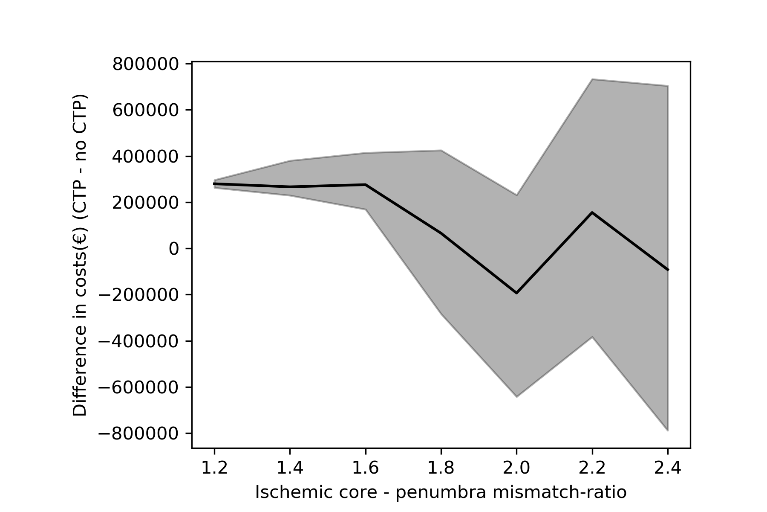

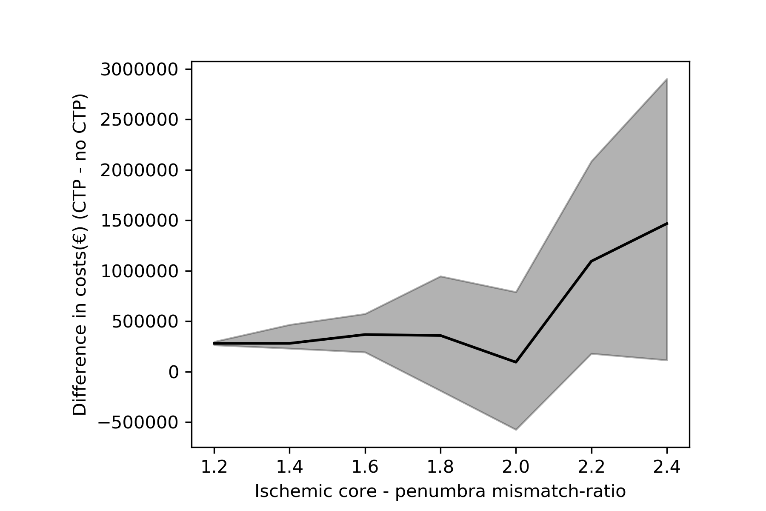

**Figure F.4: ΔCosts per core-penumbra mismatch ratio threshold per 1000 patients.** Values are presented as median with IQR for the baseline PSA. Left: 5-year follow-up. Right: 10 year follow-up.

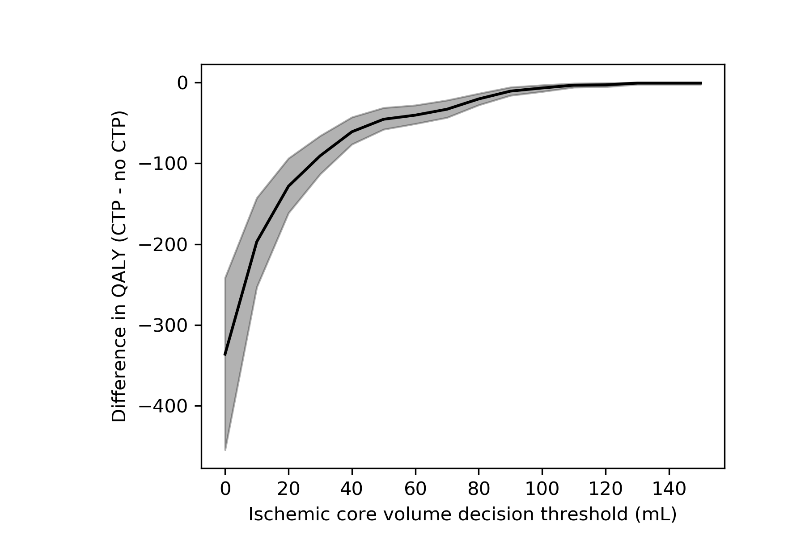

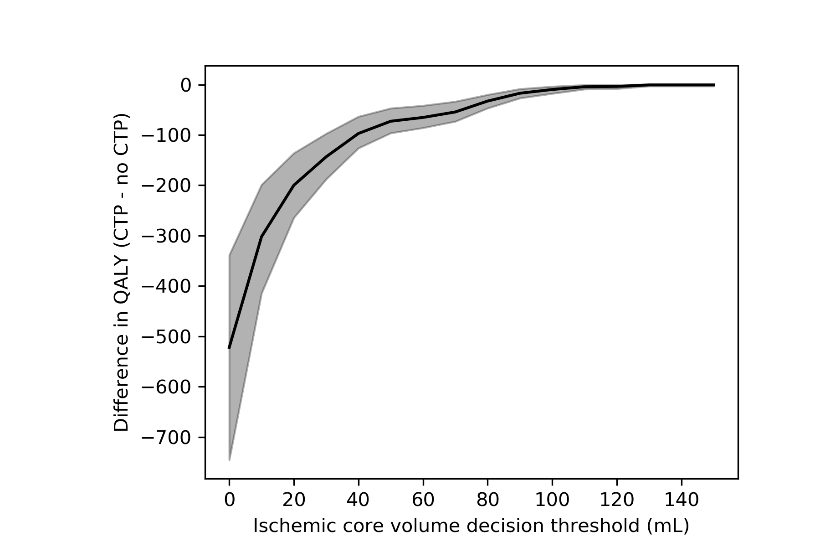

**Figure F.5: ΔQALYs per ischemic core volume decision threshold per 1000 patients.** Values are presented as median with IQR for the baseline PSA. Left: 5-year follow-up. Right: 10 year follow-up.

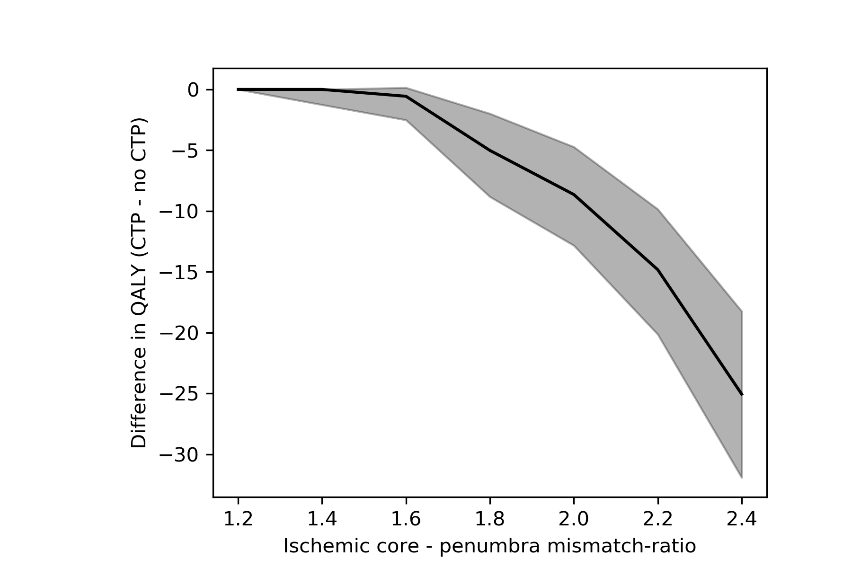

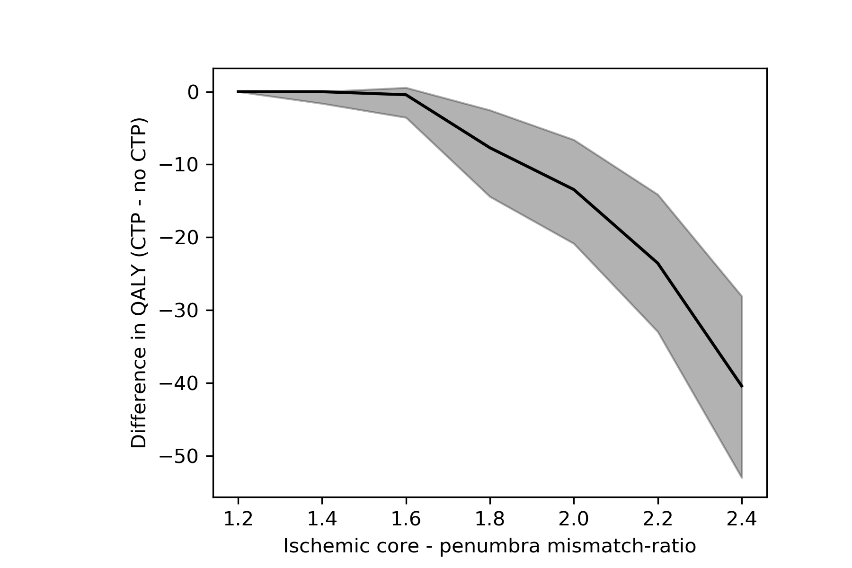

**Figure F.6: ΔQALYs per core-penumbra mismatch ratio threshold per 1000 patients.** Values are presented as median with IQR for the baseline PSA. Left: 5-year follow-up. Right: 10-year follow-up.

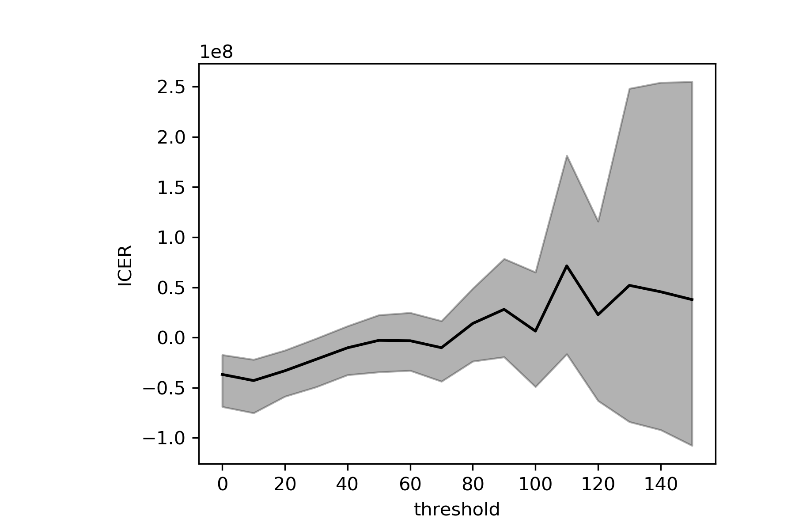

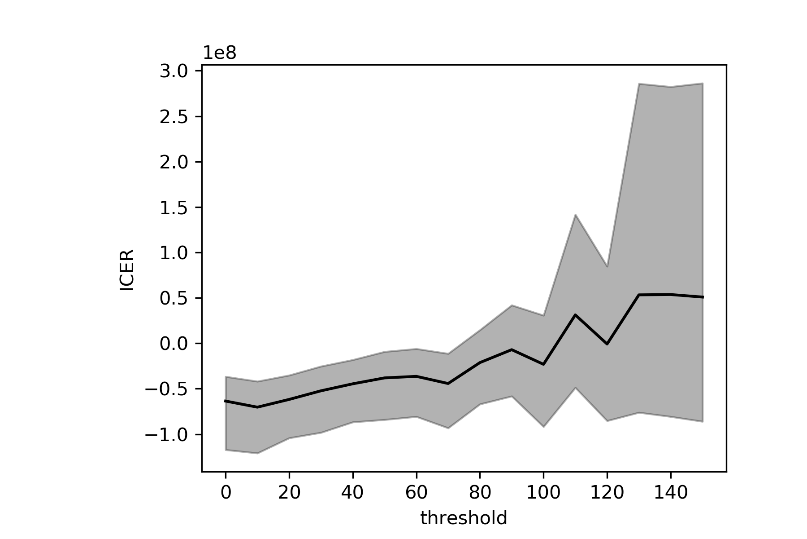

**Figure F.7: ICER per ischemic core volume threshold per 1000 patients.** Values are presented as median with IQR for the baseline PSA. Left: 5-year follow-up. Right: 10-year follow-up.

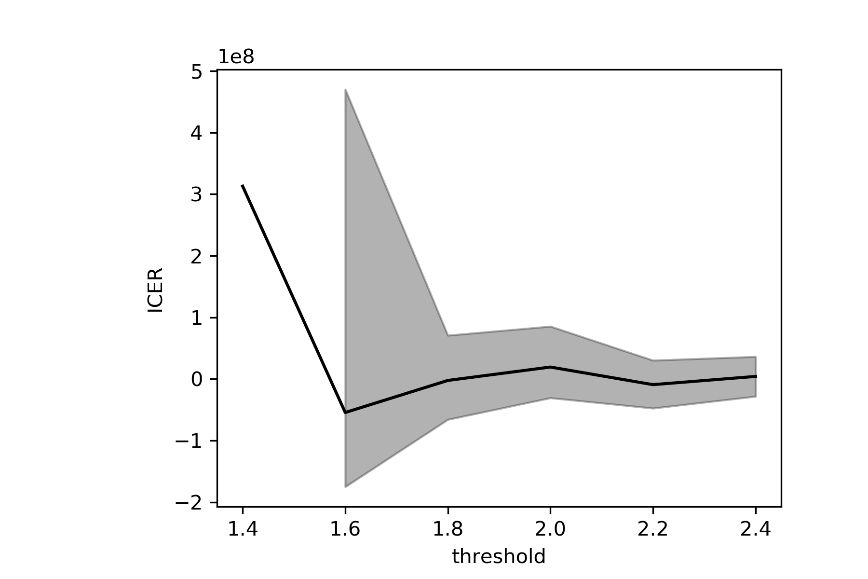

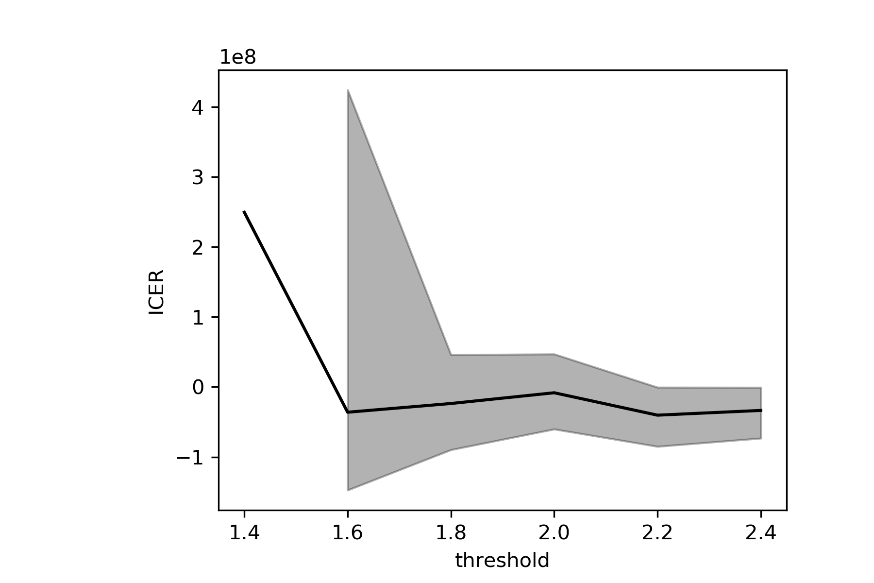

**Figure F.8: ICER per core-penumbra mismatch ratio threshold per 1000 patients.** Values are presented as median with IQR for the baseline PSA. Left: 5-year follow-up. Right: 10-year follow-up.

**Supplement G: Extended results table**

| **CTP measure** | **Years of follow-up** | **OR EVT effect (change relative to 1.67[95%CI: 1.21-2.3])** | **OR effect modification (change relative to baseline)*** | **ICER**  **Median (IQR)** | **NMB (€)**  **Median (IQR)** | **ΔQALYs**  **Median (IQR)** | **ΔCosts (€)**  **Median (IQR)** |
| --- | --- | --- | --- | --- | --- | --- | --- |
| ICV≥110mL | 5 | -0.5 | -0.1 | 57035 (-81387;213725) | 44361 (-84207;180651) | -0.576 (-1.709;0.026) | -98566 (-244442;14011) |
|  | 5 | -0.5 | -0.05 | 54204 (-52029;194974) | 52532 (-158294;247178) | -1.324 (-3.112;-0.124) | -150364 (-394145;15489) |
|  | 5 | -0.5 | 0 | 63571 (-27445;182863) | 75260 (-256606;356220) | -2.55 (-4.962;-0.677) | -262666 (-589364;-24891) |
|  | 5 | -0.4 | -0.1 | 56112 (-80919;202785) | 43761 (-91557;186341) | -0.634 (-1.82;0.01) | -103520 (-257824;14687) |
|  | 5 | -0.4 | -0.05 | 56048 (-47465;191643) | 55220 (-167300;258060) | -1.454 (-3.342;-0.17) | -162788 (-413780;14303) |
|  | 5 | -0.4 | 0 | 67394 (-22823;181803) | 84206 (-267820;369916) | -2.748 (-5.202;-0.77) | -282565 (-621968;-31396) |
|  | 5 | -0.3 | -0.1 | 56115 (-79143;200186) | 45137 (-97373;189920) | -0.697 (-1.918;-0.0) | -108139 (-272493;15091) |
|  | 5 | -0.3 | -0.05 | 56816 (-45700;187411) | 58740 (-171751;270871) | -1.584 (-3.53;-0.199) | -172610 (-434498;12927) |
|  | 5 | -0.3 | 0 | 68218 (-19427;182480) | 92540 (-274492;387168) | -2.878 (-5.369;-0.866) | -300255 (-646764;-37602) |
|  | 5 | -0.2 | -0.1 | 56679 (-75842;199414) | 47333 (-103038;198221) | -0.77 (-2.025;-0.001) | -113113 (-283348;14654) |
|  | 5 | -0.2 | -0.05 | 58023 (-42033;194116) | 61656 (-175985;281418) | -1.702 (-3.679;-0.254) | -182325 (-454905;6496) |
|  | 5 | -0.2 | 0 | 69736 (-17511;185504) | 97328 (-279658;406186) | -3.019 (-5.561;-0.943) | -317090 (-671630;-42393) |
|  | 5 | -0.1 | -0.1 | 56879 (-72084;201236) | 50367 (-108819;203738) | -0.829 (-2.136;-0.009) | -117656 (-297047;15480) |
|  | 5 | -0.1 | -0.05 | 61153 (-40076;198122) | 66147 (-177890;288937) | -1.787 (-3.829;-0.287) | -190396 (-472011;148) |
|  | 5 | -0.1 | 0 | 70378 (-16496;182958) | 100154 (-282228;416840) | -3.151 (-5.749;-0.998) | -333259 (-693926;-46221) |
|  | 5 | 0 | -0.1 | 55947 (-71851;202198) | 52380 (-117014;211993) | -0.894 (-2.251;-0.023) | -123386 (-309257;13828) |
|  | 5 | 0 | -0.05 | 63146 (-36895;198058) | 71360 (-189586;298635) | -1.867 (-3.982;-0.332) | -200018 (-490399;-1770) |
|  | 5 | 0 | 0 | 71346 (-16517;181241) | 102227 (-282942;431923) | -3.27 (-5.874;-1.052) | -348966 (-712406;-51158) |
|  | 5 | 0.1 | -0.1 | 56200 (-69553;200735) | 54241 (-124656;220749) | -0.961 (-2.349;-0.033) | -128429 (-320794;15006) |
|  | 5 | 0.1 | -0.05 | 65735 (-34476;195958) | 75914 (-193954;308028) | -1.959 (-4.127;-0.371) | -215795 (-506732;-4882) |
|  | 5 | 0.1 | 0 | 73472 (-12106;182197) | 105745 (-283315;447098) | -3.38 (-6.055;-1.102) | -366110 (-725442;-54417) |
|  | 5 | 0.2 | -0.1 | 57846 (-67800;200167) | 55301 (-128324;225883) | -1.022 (-2.449;-0.043) | -134942 (-331458;14009) |
|  | 5 | 0.2 | -0.05 | 67269 (-31691;194190) | 77932 (-195506;318772) | -2.055 (-4.28;-0.404) | -226367 (-520125;-7100) |
|  | 5 | 0.2 | 0 | 74175 (-10997;181061) | 107731 (-281118;460564) | -3.502 (-6.225;-1.146) | -378798 (-740749;-58667) |
|  | 5 | 0.3 | -0.1 | 57417 (-66942;196961) | 57805 (-131228;233197) | -1.085 (-2.556;-0.05) | -139714 (-344263;14650) |
|  | 5 | 0.3 | -0.05 | 65110 (-31006;189307) | 81540 (-202099;334318) | -2.154 (-4.414;-0.454) | -232732 (-535444;-8026) |
|  | 5 | 0.3 | 0 | 75009 (-10030;180919) | 115014 (-282947;472304) | -3.624 (-6.366;-1.188) | -393648 (-754846;-64458) |
|  | 5 | 0.82 | -0.1 | 58939 (-57998;197632) | 69596 (-146628;265746) | -1.354 (-3.068;-0.099) | -158741 (-397187;9673) |
|  | 5 | 0.82 | -0.05 | 72948 (-19370;189290) | 95139 (-235410;378627) | -2.588 (-5.031;-0.654) | -284265 (-621398;-23967) |
|  | 5 | 0.82 | 0 | 79288 (-3228;187236) | 126436 (-294179;516591) | -4.0 (-6.968;-1.43) | -436485 (-823877;-86247) |
|  | 10 | -0.5 | -0.1 | 13924 (-107627;147588) | -24078 (-262408;153870) | -0.174 (-1.797;1.033) | 8203 (-149320;225180) |
|  | 10 | -0.5 | -0.05 | 15434 (-95929;151501) | -82733 (-430820;215513) | -0.994 (-3.999;0.927) | -14184 (-291235;270693) |
|  | 10 | -0.5 | 0 | 22562 (-68585;136198) | -119896 (-587168;333612) | -2.981 (-7.277;-0.044) | -145266 (-568643;244701) |
|  | 10 | -0.4 | -0.1 | 14070 (-110030;145371) | -30516 (-281916;157278) | -0.225 (-1.947;1.015) | 8908 (-156743;226729) |
|  | 10 | -0.4 | -0.05 | 14205 (-97628;144677) | -85256 (-447245;233845) | -1.133 (-4.292;0.867) | -24232 (-316069;269778) |
|  | 10 | -0.4 | 0 | 23605 (-66187;137205) | -119281 (-603941;345315) | -3.309 (-7.651;-0.159) | -167871 (-589762;239434) |
|  | 10 | -0.3 | -0.1 | 15407 (-107006;150976) | -38411 (-295091;163770) | -0.278 (-2.135;1.022) | 8637 (-166152;237296) |
|  | 10 | -0.3 | -0.05 | 16365 (-91996;145559) | -89848 (-460125;247493) | -1.297 (-4.514;0.728) | -36184 (-338417;262836) |
|  | 10 | -0.3 | 0 | 25893 (-60577;139203) | -112504 (-620679;366957) | -3.537 (-7.982;-0.262) | -186259 (-618667;225119) |
|  | 10 | -0.2 | -0.1 | 17228 (-106133;153708) | -42180 (-307595;172895) | -0.331 (-2.308;1.035) | 4985 (-174070;240083) |
|  | 10 | -0.2 | -0.05 | 15346 (-92066;137154) | -95364 (-476997;258349) | -1.465 (-4.817;0.664) | -43509 (-360974;259447) |
|  | 10 | -0.2 | 0 | 27653 (-55784;139526) | -112212 (-626647;382943) | -3.811 (-8.364;-0.363) | -209420 (-653387;214308) |
|  | 10 | -0.1 | -0.1 | 15957 (-107522;149440) | -44065 (-317094;179724) | -0.422 (-2.488;1.012) | 2557 (-189977;245799) |
|  | 10 | -0.1 | -0.05 | 18261 (-90217;139386) | -95683 (-492351;262064) | -1.633 (-4.989;0.618) | -60346 (-382490;258983) |
|  | 10 | -0.1 | 0 | 29231 (-53582;138694) | -104584 (-638723;405304) | -4.097 (-8.698;-0.535) | -228620 (-690705;204550) |
|  | 10 | 0 | -0.1 | 16219 (-102805;158664) | -47193 (-327253;184168) | -0.499 (-2.652;1.017) | -3120 (-203254;249522) |
|  | 10 | 0 | -0.05 | 18948 (-88290;138981) | -95305 (-508231;268169) | -1.773 (-5.295;0.527) | -68605 (-407677;258424) |
|  | 10 | 0 | 0 | 31244 (-48780;141749) | -97781 (-644889;420136) | -4.346 (-9.065;-0.651) | -246679 (-733714;191840) |
|  | 10 | 0.1 | -0.1 | 17184 (-99660;163819) | -51098 (-344249;189337) | -0.583 (-2.793;1.008) | -5263 (-215647;247064) |
|  | 10 | 0.1 | -0.05 | 19639 (-82503;140040) | -96881 (-507471;275083) | -2.018 (-5.559;0.386) | -81194 (-436340;259133) |
|  | 10 | 0.1 | 0 | 32885 (-47053;141486) | -101056 (-650787;437771) | -4.57 (-9.35;-0.772) | -269124 (-774042;181113) |
|  | 10 | 0.2 | -0.1 | 16340 (-103000;159953) | -54225 (-357808;198961) | -0.659 (-2.958;0.995) | -8581 (-234230;252198) |
|  | 10 | 0.2 | -0.05 | 21703 (-81096;141291) | -98816 (-513571;288845) | -2.183 (-5.814;0.318) | -93835 (-453393;252877) |
|  | 10 | 0.2 | 0 | 34370 (-44782;141426) | -105302 (-654179;447107) | -4.801 (-9.596;-0.88) | -280358 (-806374;169203) |
|  | 10 | 0.3 | -0.1 | 16328 (-101862;158015) | -56497 (-369169;205562) | -0.737 (-3.073;0.98) | -10532 (-253472;253672) |
|  | 10 | 0.3 | -0.05 | 23106 (-76081;144383) | -94125 (-521341;297295) | -2.323 (-6.048;0.222) | -103309 (-475934;249412) |
|  | 10 | 0.3 | 0 | 35650 (-44191;141302) | -103484 (-659676;460891) | -5.018 (-9.771;-0.999) | -295151 (-832410;151728) |
|  | 10 | 0.82 | -0.1 | 15440 (-103939;153179) | -69079 (-415466;234382) | -1.061 (-3.901;0.865) | -32815 (-315893;259125) |
|  | 10 | 0.82 | -0.05 | 25928 (-65763;137779) | -96801 (-565620;347538) | -3.099 (-7.207;-0.016) | -160655 (-571570;220386) |
|  | 10 | 0.82 | 0 | 39788 (-38956;141756) | -74712 (-685404;519196) | -5.824 (-10.778;-1.566) | -373317 (-951864;101952) |
| MMR≤1.4 | 5 | -0.5 | 0 | 380124 (-151368;-2147483648) | -263889 (-433414;-222141) | 0.0 (-0.976;0.0) | 272937 (229403;377940) |
|  | 5 | -0.5 | 0.05 | 380566 (-151071;-2147483648) | -264070 (-433655;-222185) | 0.0 (-0.98;0.0) | 272851 (229403;377990) |
|  | 5 | -0.5 | 0.1 | 380991 (-150789;-2147483648) | -264073 (-433887;-222227) | 0.0 (-0.984;0.0) | 272766 (229403;378038) |
|  | 5 | -0.4 | 0 | 361345 (-149258;-2147483648) | -264242 (-438308;-223070) | 0.0 (-1.053;0.0) | 271425 (229403;379180) |
|  | 5 | -0.4 | 0.05 | 361774 (-149006;-2147483648) | -264427 (-438604;-223111) | 0.0 (-1.056;0.0) | 271334 (229403;379217) |
|  | 5 | -0.4 | 0.1 | 362187 (-148728;-2147483648) | -264604 (-438887;-223151) | 0.0 (-1.058;0.0) | 271247 (229403;379252) |
|  | 5 | -0.3 | 0 | 349751 (-145069;-2147483648) | -267393 (-443666;-225803) | 0.0 (-1.092;0.0) | 269980 (229403;379943) |
|  | 5 | -0.3 | 0.05 | 350047 (-144842;-2147483648) | -267548 (-444090;-225943) | 0.0 (-1.094;0.0) | 269887 (229403;379994) |
|  | 5 | -0.3 | 0.1 | 350331 (-144625;-2147483648) | -267696 (-444428;-226076) | 0.0 (-1.096;0.0) | 269798 (229403;380042) |
|  | 5 | -0.2 | 0 | 330581 (-143474;-2147483648) | -270117 (-449671;-225297) | 0.0 (-1.152;0.0) | 268606 (229403;380813) |
|  | 5 | -0.2 | 0.05 | 330819 (-143169;-2147483648) | -270291 (-449886;-225155) | 0.0 (-1.156;0.0) | 268513 (229403;380856) |
|  | 5 | -0.2 | 0.1 | 331047 (-142878;-2147483648) | -270600 (-450092;-225020) | 0.0 (-1.16;0.0) | 268423 (229403;380881) |
|  | 5 | -0.1 | 0 | 311909 (-143710;-2147483648) | -276027 (-453103;-227992) | 0.0 (-1.216;0.0) | 267509 (229403;380790) |
|  | 5 | -0.1 | 0.05 | 312431 (-143469;-2147483648) | -276354 (-453405;-228199) | 0.0 (-1.219;0.0) | 267440 (229403;380809) |
|  | 5 | -0.1 | 0.1 | 312935 (-143240;-2147483648) | -276667 (-453692;-228398) | 0.0 (-1.223;0.0) | 267374 (229403;380827) |
|  | 5 | 0 | 0 | 312955 (-141379;-2147483648) | -278626 (-456559;-229403) | 0.0 (-1.261;0.0) | 266513 (229403;380110) |
|  | 5 | 0 | 0.05 | 301586 (-145043;-2147483648) | -278740 (-456835;-229403) | 0.0 (-1.264;0.0) | 266426 (229403;380103) |
|  | 5 | 0 | 0.1 | 301902 (-144785;-2147483648) | -278850 (-457097;-229403) | 0.0 (-1.267;0.0) | 266336 (229403;380095) |
|  | 5 | 0.1 | 0 | 296658 (-143052;-2147483648) | -279614 (-459631;-229403) | 0.0 (-1.318;0.0) | 265626 (229403;380329) |
|  | 5 | 0.1 | 0.05 | 297041 (-142699;-2147483648) | -279614 (-459707;-229403) | 0.0 (-1.323;0.0) | 265592 (229403;380375) |
|  | 5 | 0.1 | 0.1 | 297610 (-142370;-2147483648) | -279614 (-459779;-229403) | 0.0 (-1.328;0.0) | 265556 (229403;380396) |
|  | 5 | 0.2 | 0 | 305773 (-138869;-2147483648) | -279614 (-462068;-229403) | 0.0 (-1.373;0.0) | 265251 (229403;380989) |
|  | 5 | 0.2 | 0.05 | 306263 (-138108;-2147483648) | -279614 (-462412;-229403) | 0.0 (-1.377;0.0) | 265248 (229403;381054) |
|  | 5 | 0.2 | 0.1 | 306733 (-137924;-2147483648) | -279614 (-462741;-229403) | 0.0 (-1.38;0.0) | 265245 (229403;381116) |
|  | 5 | 0.3 | 0 | 296185 (-141271;-2147483648) | -279614 (-465361;-229379) | 0.0 (-1.41;0.0) | 265066 (229403;381391) |
|  | 5 | 0.3 | 0.05 | 296456 (-141035;-2147483648) | -279614 (-465472;-229360) | 0.0 (-1.414;0.0) | 265039 (229403;381437) |
|  | 5 | 0.3 | 0.1 | 296647 (-140811;-2147483648) | -279614 (-465577;-229342) | 0.0 (-1.418;0.0) | 265013 (229403;381481) |
|  | 5 | 0.82 | 0 | 287382 (-137089;-2147483648) | -279614 (-477472;-229403) | 0.0 (-1.551;0.0) | 262877 (229374;382923) |
|  | 5 | 0.82 | 0.05 | 286035 (-137386;-2147483648) | -279614 (-477681;-229403) | 0.0 (-1.553;0.0) | 262877 (229354;382937) |
|  | 5 | 0.82 | 0.1 | 286266 (-137225;-2147483648) | -279614 (-477880;-229403) | 0.0 (-1.555;0.0) | 262877 (229345;382949) |
|  | 10 | -0.5 | 0 | 331325 (-110302;-2147483648) | -271910 (-520757;-219184) | 0.0 (-1.038;0.307) | 295628 (229403;466475) |
|  | 10 | -0.5 | 0.05 | 327882 (-113245;-2147483648) | -272027 (-521338;-219251) | 0.0 (-1.044;0.302) | 295569 (229403;466505) |
|  | 10 | -0.5 | 0.1 | 328710 (-112631;-2147483648) | -272133 (-521893;-219316) | 0.0 (-1.05;0.296) | 295454 (229403;466534) |
|  | 10 | -0.4 | 0 | 322138 (-114173;-2147483648) | -274968 (-528942;-221327) | 0.0 (-1.127;0.196) | 292704 (229403;466808) |
|  | 10 | -0.4 | 0.05 | 319479 (-116564;-2147483648) | -275462 (-529312;-221356) | 0.0 (-1.13;0.195) | 292593 (229403;466793) |
|  | 10 | -0.4 | 0.1 | 319956 (-116191;-2147483648) | -275934 (-529665;-221433) | 0.0 (-1.135;0.193) | 292444 (229403;466778) |
|  | 10 | -0.3 | 0 | 293186 (-122265;-2147483648) | -279614 (-534853;-222380) | 0.0 (-1.246;0.141) | 288922 (229403;466738) |
|  | 10 | -0.3 | 0.05 | 293938 (-121983;-2147483648) | -279614 (-535334;-222371) | 0.0 (-1.253;0.136) | 288772 (229403;466768) |
|  | 10 | -0.3 | 0.1 | 294662 (-121715;-2147483648) | -279614 (-535771;-222362) | 0.0 (-1.26;0.132) | 288628 (229403;466795) |
|  | 10 | -0.2 | 0 | 304010 (-117442;-2147483648) | -279614 (-542881;-224284) | 0.0 (-1.382;0.08) | 285081 (229403;466778) |
|  | 10 | -0.2 | 0.05 | 303051 (-117788;-2147483648) | -279614 (-543347;-224485) | 0.0 (-1.388;0.076) | 284647 (229403;466766) |
|  | 10 | -0.2 | 0.1 | 304298 (-117523;-2147483648) | -279614 (-543747;-224678) | 0.0 (-1.393;0.073) | 284232 (229403;466755) |
|  | 10 | -0.1 | 0 | 273173 (-120297;-2147483648) | -279614 (-548826;-227168) | 0.0 (-1.495;0.0) | 281253 (229403;466543) |
|  | 10 | -0.1 | 0.05 | 273727 (-120081;-2147483648) | -279614 (-549431;-227366) | 0.0 (-1.503;0.0) | 281032 (229403;466508) |
|  | 10 | -0.1 | 0.1 | 274168 (-119875;-2147483648) | -279614 (-549938;-227556) | 0.0 (-1.509;0.0) | 280821 (229403;466475) |
|  | 10 | 0 | 0 | 249523 (-118325;-2147483648) | -279614 (-555168;-228836) | 0.0 (-1.619;0.0) | 279661 (229403;464756) |
|  | 10 | 0 | 0.05 | 249728 (-118011;-2147483648) | -279614 (-555585;-228917) | 0.0 (-1.624;0.0) | 279632 (229403;464830) |
|  | 10 | 0 | 0.1 | 248872 (-118899;-2147483648) | -279614 (-555982;-228996) | 0.0 (-1.629;0.0) | 279614 (229403;464901) |
|  | 10 | 0.1 | 0 | 245092 (-118855;-2147483648) | -279614 (-562241;-229403) | 0.0 (-1.699;0.0) | 279614 (229403;464310) |
|  | 10 | 0.1 | 0.05 | 244511 (-120063;-2147483648) | -279614 (-562893;-229403) | 0.0 (-1.706;0.0) | 279614 (229403;464325) |
|  | 10 | 0.1 | 0.1 | 244943 (-119699;-2147483648) | -279614 (-563408;-229403) | 0.0 (-1.712;0.0) | 279614 (229403;464325) |
|  | 10 | 0.2 | 0 | 242770 (-118070;-2147483648) | -279614 (-566573;-229401) | 0.0 (-1.804;0.0) | 279614 (229403;463657) |
|  | 10 | 0.2 | 0.05 | 242140 (-119736;-2147483648) | -279614 (-566742;-229403) | 0.0 (-1.81;0.0) | 279614 (229403;463724) |
|  | 10 | 0.2 | 0.1 | 242401 (-119449;-2147483648) | -279614 (-566913;-229403) | 0.0 (-1.815;0.0) | 279614 (229403;463788) |
|  | 10 | 0.3 | 0 | 240182 (-117772;-2147483648) | -279614 (-570464;-229403) | 0.0 (-1.865;0.0) | 279614 (229403;464187) |
|  | 10 | 0.3 | 0.05 | 240481 (-117346;-2147483648) | -279614 (-570770;-229403) | 0.0 (-1.87;0.0) | 279614 (229403;464165) |
|  | 10 | 0.3 | 0.1 | 240768 (-117138;-2147483648) | -279614 (-571036;-229403) | 0.0 (-1.878;0.0) | 279614 (229403;464143) |
|  | 10 | 0.82 | 0 | 229615 (-110087;-2147483648) | -279614 (-589096;-229403) | 0.0 (-2.198;0.0) | 279614 (229403;460911) |
|  | 10 | 0.82 | 0.05 | 229761 (-109921;-2147483648) | -279614 (-589487;-229403) | 0.0 (-2.202;0.0) | 279614 (229403;460825) |
|  | 10 | 0.82 | 0.1 | 229902 (-109763;-2147483648) | -279614 (-589859;-229403) | 0.0 (-2.207;0.0) | 279614 (229403;460750) |

**Table G1. Extended results table per 1000 patients.** An extended version of Table 2 in the manuscript. Results are presented as median with IQR, the baseline follow-up horizon of 5-year was used. For all scenarios, the optimal ICV threshold in was 110mL and the optimal MMR threshold was 1.4. Optimal thresholds were based on the maximum net monetary benefit at a willingness to pay of €80000. *: Baseline EVT effect modification ORs were 0.98 (95%CI:0.881;1.091) for ICV and 1.010 (95%CI:0.994;1.560) for MMR. ICER: incremental cost effectiveness ratio. ICV: ischaemic core volume. MMR: core-penumbra mismatch ratio. EVT: endovascular treatment. OR: odds ratio. CTP: CT perfusion.

**Online supplement H. Effects in included sub-populations**

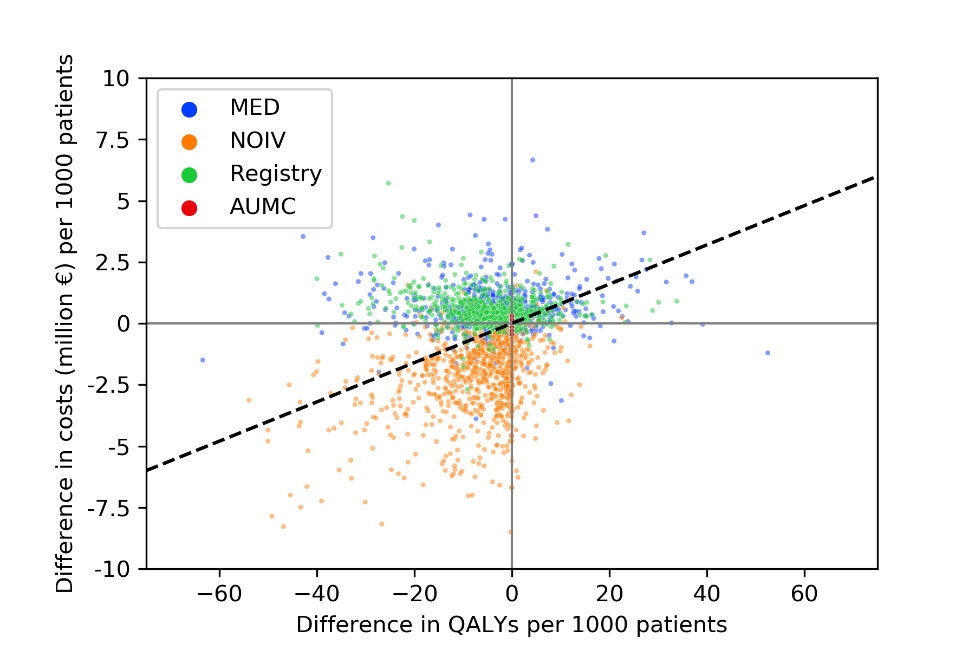

**Figure H1. Variations in outcome per data source.** The optimal ischemic core volume threshold of ≥110mL was used to exclude patients for EVT. Regardless of the data source used, the conclusion was the same. For patients from the MR CLEAN NOIV data, included within 4.5 hours after symptom onset due to intravenous thrombolysis eligibility criteria for inclusion, the cost savings were more profound. This was attributed to a shorter survival (and less costs) of patients with a relatively good baseline 90-day mRS outcome if patients were excluded for EVT.

**Supplementary material references**

9.	Dutch Central Bureau of Statistics. Consumer price index [Internet]. 2020 [cited 2020 Sep 1]. Available from: opendata.cbs.nl/statline

10.	Van Den Berg LA, Berkhemer OA, Fransen PSS, Beumer D, Lingsma H, Majoie CBM, et al. Economic Evaluation of Endovascular Treatment for Acute Ischemic Stroke. Stroke. 2022;29(2):968–75.

11.	Van Voorst H, Kunz WG, Van Den Berg LA, Kappelhof M, Pinckaers FME, Goyal M, et al. Quantified health and cost effects of faster endovascular treatment for large vessel ischemic stroke patients in the Netherlands. J Neurointerv Surg. 2020;1–8.

12.	Hakkaart-van Roijen L, van der Linden N, Bouwmans C, Kanters T, Swan Tan S. Kostenhandleiding: Methodologie van kostenonderzoek en referentieprijzen voor economische evaluaties in de gezondheidszorg. Dutch Natl Heal Care Inst. 2016;

13.	Van Voorst H, Kunz WG, Van Den Berg LA, Kappelhof M, Pinckaers FME, Goyal M, et al. Quantified health and cost effects of faster endovascular treatment for large vessel ischemic stroke patients in the Netherlands. J Neurointerv Surg. 2021;13(12):1099–105.
